# Development and multi-dataset evaluation of a unified single-view deep-learning model for the right heart: four-chamber segmentation, biventricular ejection fraction, deformation, and pulmonary-hypertension prediction from the apical four-chamber echocardiogram

**DOI:** 10.64898/2026.08.19.26360852

**Authors:** Tyler Pitre, Lais Marques, Jason Weatherald, Susanna Mak, Paaladinesh Thavendiranathan, John Granton

**Affiliations:** Pulmonary Hypertension Programme, Toronto General Hospital, University Health Network, Toronto, ON, Canada; Division of Cardiology, Department of Medicine, Schulich School of Medicine, Western University, London, ON, Canada; Division of Pulmonary Medicine, Lung Transplant & Pulmonary Hypertension, University of, Alberta & Alberta Health Services; Division of Cardiology, Mount Sinai Hospital, Sinai Health System Ted Rogers Program in Cardiotoxicity Prevention & Peter Munk Cardiac Centre, Toronto General Hospital, University

**Keywords:** echocardiography, right ventricle, deep learning, ejection fraction, myocardial strain, pulmonary hypertension, multi-task learning, external validation, TRIPOD+AI

## Abstract

**Background:** Right ventricular (RV) function predicts survival in pulmonary hypertension (PH) and other cardiovascular diseases, yet echocardiographic AI has largely focused on the left ventricle (LV).

**Objectives:** To develop and evaluate PH-ECHO-AI, a unified deep learning model performing four-chamber segmentation, landmark localisation, biventricular ejection fraction (EF) estimation, deformation analysis, and PH prediction from a single apical four-chamber (A4C) clip.

**Methods:** We developed the model using 8,416 clips from four public datasets and no institutional data: EchoNet-Dynamic, CAMUS, RVENet (apical four-chamber clips paired with 3D-echocardiographic right ventricular ejection fraction, RVEF), and MIMIC-IV-ECHO. Evaluation used held-out, training-excluded data with expert-reviewed reference standards and a per-cohort audit of patient-level separation: 1,416 clips for segmentation; 600 clips for function and deformation (350 referenced to 3D-echocardiographic RVEF, 250 to the EchoNet LVEF); and 1,076 MIMIC-IV patients for PH prediction, with five-fold cross-validation. Performance measures were Dice, correlation, mean absolute error (MAE), Bland-Altman agreement, and area under the receiver operating characteristic curve (AUC).

**Results:** Four-chamber segmentation generalised robustly across all datasets (pooled Dice: LV 0.925, RV 0.836, LA 0.910, RA 0.904). Left ventricular ejection fraction (LVEF) was estimated with r=0.845 (95% CI 0.786 to 0.886) and MAE 4.67%. RVEF, regressed directly from the clip by a supervised head trained on 3D-echocardiographic labels with no geometric assumption, reached r=0.754 (95% CI 0.690 to 0.806) and MAE 4.98%, matching published single-view RVEF ceilings and exceeding geometric RV fractional area change (RVFAC; r=0.278). Deformation and excursion metrics, namely RV free-wall and LV A4C longitudinal strain and tricuspid and mitral annular plane systolic excursion (TAPSE, MAPSE), proved physiologically coherent. Segmentation generalised to the external MIMIC-IV cohort, and PH prediction was developed and evaluated entirely within it; RVEF evaluation was clip-disjoint and same-source, so cross-centre RVEF validation remains outstanding. Using echocardiographic geometry alone, confirmed PH was detected with an AUC of 0.697 and strong calibration (Brier 0.061).

**Conclusions:** A single, reproducible model provides comprehensive right-heart-focused interpretation from one A4C view. It achieves RVEF accuracy competitive with dedicated RV models while simultaneously delivering segmentation, deformation, annular excursion (TAPSE and MAPSE), and PH prediction.

**Registration:** This retrospective study used existing datasets. Code is openly released, and trained model weights are available to credentialed investigators, for independent evaluation.

## Introduction

Right ventricular (RV) function is a dominant determinant of outcome across cardiovascular disease and is the principal mode of death in pulmonary hypertension (PH) (1). Yet the right ventricle is the least reliably quantified structure in routine echocardiography. This is in part due to its crescentic geometry, retrosternal position, and load dependence making manual assessment difficult and poorly reproducible (1, 2). Furthermore, three-dimensional (3D) RV ejection fraction (RVEF), the most prognostically informative index, which outperforms conventional measures for predicting mortality (3), requires dedicated acquisition and offline software unavailable in most laboratories and infeasible at the point of care.

Artificial intelligence has transformed left-ventricle quantification. EchoNet-Dynamic achieved expert-level LV ejection fraction (LVEF) estimation and LV segmentation from a single video (4), later shown in a blinded randomised trial to be superior to sonographer assessment in clinical workflow (5). Foundational models have extended automated interpretation to many tasks (6, 7). The automated quantification of right ventricular function is more limited. The most advanced dedicated tool, EchoNet-RV, segments the RV and estimates RV fractional area change (RVFAC), a two-dimensional (2D) surrogate, reporting superior right-heart performance to general foundation models (8); a separate line of work estimates RVEF directly from 2D video against 3D-echo ground truth (9). Each existing approach is single-purpose: a complete right-ventricle read still requires assembling separate segmentation, function, deformation, and view-classification models.

Access to echocardiography, and to specialised RV assessment, is unequally distributed; under-resourced and point-of-care settings are precisely those least likely to have 3D RV software. We also note that the development data are drawn predominantly from single academic centres in high-income countries, which may not represent all sociodemographic groups.

We aimed (i) to develop a single multi-task model that performs four-chamber segmentation, landmark localisation, direct biventricular EF estimation, deformation/excursion quantification, and PH prediction from one A4C clip; (ii) to evaluate each capability on held-out, training-excluded data with expert reference standards, including RVEF against 3D echocardiography, auditing patient-level separation per cohort rather than assuming it; and (iii) to benchmark performance against the published state of the art. The study therefore reports both development and evaluation of prediction models (the segmentation network is the upstream representation on which the EF and PH prediction models depend).

## Methods

### Source of data and study design

This retrospective study developed and evaluated prediction models on four public echocardiographic datasets selected for complementary strengths (Supplementary Table S1): EchoNet-Dynamic (Stanford; reference LVEF and expert LV tracings) for the LV reference and large-scale A4C training; CAMUS (expert LV/LA masks augmented with investigator four-chamber masks) for multi-chamber ground truth; RVENet (Semmelweis; 2D A4C DICOM with 3D-echo-derived RVEF) for the RVEF reference; and MIMIC-IV-ECHO (Beth Israel Deaconess; routine clinical clips with administrative, text, RVSP and catheter PH labels) for external generalisation and the PH cohort. Acquisition windows appear in Supplementary Table S1. The datasets are convenience samples rather than population-representative cohorts.

We report this manuscript in accordance with the TRIPOD+AI checklist (10).

### Participants, setting, and treatments

Each dataset originates from a single centre; together they span four centres across three countries and two acquisition conventions (standard A4C with the RV on the image left; Mayo convention with the RV on the right). Eligible clips were A4C acquisitions with interpretable cardiac anatomy; clips failing automated quality control (below) were excluded from measurement and from the PH cohort. For the PH cohort, MIMIC-IV patients with linkable labels were eligible. Treatments were not used as model inputs and were not part of outcome definitions; this is a cross-sectional measurement and diagnostic-prediction study, so treatment handling is not applicable.

### Data preparation and quality control

DICOM and video clips underwent luminance extraction, resizing to 224×224 (model input) and 256×256 (measurement space), and per-frame intensity normalisation; the same preprocessing was applied uniformly across all datasets and subgroups. Acquisition orientation was handled natively by the model; for the segmentation Dice evaluation only (11), a mask-derived 90°-rotation/mirror canonicalisation was applied identically to image and label so stored and canonical orientations could be compared fairly. Every evaluation clip was excluded from training by identifier, leaving 8,416 clips for training. Each measurement carries a model-native quality score (mask-boundary fidelity, frame stability, segment survival, landmark confidence) with reportable, caution and not-reportable tiers; its distribution appears in Supplementary Table S2.

### Outcomes and reference standards

The prediction tasks, their outcomes, reference standards, and decision thresholds are summarised in Supplementary Table S3. The EF and PH outcomes are concurrent/diagnostic, so no prognostic time horizon applies. Reference LVEF and RVEF were generated independently of our model. Reference LVEF is the EchoNet-Dynamic value: expert apical four-chamber tracings integrated by the single-plane method of disks (the dataset releases only that view), obtained by a registered sonographer and verified by a level-3 echocardiographer. Reference RVEF is the RVENet 3D-echocardiographic value, which incorporates the outflow-tract contribution the four-chamber view cannot image. Cardiac magnetic resonance (CMR), the definitive reference for right-ventricular volumes, was not available in any dataset. No institutional data were used. Confirmed PH was defined as the presence of PH support (administrative codes, discharge-text mention, or PH-specific medication) together with an objective threshold, echocardiographic RV systolic pressure (RVSP) ≥40 mmHg or catheter-derived pulmonary artery systolic pressure ≥35 mmHg; controls had no PH support. Segmentation reference masks were reviewed by a domain expert. Reference standards were established without reference to model predictions, and reference EF and PH labels are pre-existing dataset annotations.

### Predictors / model inputs

For segmentation, landmark localisation, and EF, the model input (“predictor”) is the raw A4C video; no manual feature pre-selection was performed (the model is end-to-end). For the PH prediction model, eleven echocardiographic-geometric features derived by PH-ECHO-AI were used: LVEF, RVEF, RVFAC, RV free-wall longitudinal strain, LV apical four-chamber longitudinal strain, RV/LV area ratio, RV end-diastolic and end-systolic areas, LV end-diastolic area, tricuspid annular plane systolic excursion (TAPSE), and mitral annular plane systolic excursion (MAPSE). These features are produced automatically from the masks and landmarks and require no manual measurement; their assessment does not depend on subjective human interpretation at inference.

### Sample size

No a priori sample-size calculation was performed, as is typical for deep-learning development; study size was determined by available data (1,416 segmentation clips; 250 LVEF and 350 RVEF clips; 1,076 patients with 72 confirmed-PH events). Segmentation and EF estimates carry narrow CIs; the PH model, at roughly 6.5 events per variable across eleven features, is framed as proof of concept (Limitations).

### Missing data

Clips for which video could not be decoded or from which measurements could not be extracted were excluded with reasons logged. For the PH model, missing per-patient features were median-imputed within the logistic-regression pipeline; the gradient-boosted model handled missing values natively. Clips failing the quality threshold were excluded before patient-level aggregation.

### Analytical methods: model development and evaluation

Data were partitioned at the patient level into training and held-out evaluation; evaluation clips were excluded from training by identifier. EF targets were z-standardised (EF = z × 15 + 50); PH features were standardised for logistic regression and used untransformed for gradient boosting. The model (described below) was trained with a staged curriculum and an exponential-moving-average (EMA) checkpoint (12); internal validation of the measurement model used the training-excluded held-out sets. The PH model used patient-level five-fold stratified cross-validation. Performance measures were: Dice with 95% bootstrap CIs (segmentation) (11); Pearson and Spearman correlation, MAE, bias, and Bland–Altman 95% limits of agreement against reference EF (13); and AUC, Youden-point sensitivity/specificity, and Brier score with reliability diagrams (PH). All confidence intervals are bias-corrected and accelerated bootstrap intervals from 10,000 resamples of contributing patients rather than clips (Supplementary Note 3). Model updating consisted of post-hoc Platt recalibration for the PH model. Inference detail and the deformation audit corrections appear in Supplementary Note 1, with measurement robustness in Supplementary Table S4. The released code and weights reproduce all predictions. No heterogeneity model across centres was fitted; between-dataset performance is reported descriptively.

### Class imbalance

The PH outcome is imbalanced (72/1,076 ≈ 6.7% positive). Logistic regression used class weighting to preserve discrimination under imbalance; because class weighting inflates predicted probabilities, a one-parameter Platt sigmoid was then fitted (within nested cross-validation) to restore calibration to the true base rate, reducing the Brier score 3.5-fold while preserving AUC.

### Fairness

Sociodemographic metadata were limited and inconsistently available across datasets, precluding a complete fairness audit (e.g., by race/ethnicity). To partially address differential performance, RV function accuracy was examined across clinically meaningful subgroups, RVENet patient groups (athletes, healthy volunteers, heart failure with reduced ejection fraction, valvular heart disease, transplant, non-compaction), acquisition-quality grades, and standard versus RV-focused A4C views (Supplementary Table S5). A complete sociodemographic fairness evaluation is identified as required future work (Limitations).

### Model output and decision thresholds

Outputs are per-pixel five-class segmentation; seven landmark coordinates; continuous LVEF and RVEF (%); continuous deformation (%) and annular excursion (mm); and, for PH, a calibrated probability. Decision thresholds follow the ASE/EACVI chamber-quantification recommendations (2) — lower limit of normal LVEF 52% in men and 54% in women, RVEF 45%, RVFAC 35%, TAPSE 17 mm, MAPSE 11 mm — and the prognostic literature for RV free-wall strain (20). Sex is unavailable in EchoNet-Dynamic and inconsistent elsewhere, so reporting used a sex-neutral threshold: LV dysfunction, LVEF<50%; where sex is recorded, the sex-specific guideline values should be substituted. The remaining thresholds were RV dysfunction RVEF<45% and RVFAC<35%, abnormal LV A4C strain worse (less negative) than −18%, abnormal RV free-wall strain worse (less negative) than −20%, TAPSE<17 mm, MAPSE<11 mm, RV/LV area ratio ≥0.6. The PH operating point was the cross-validated Youden threshold (probability 0.63 for the uncalibrated model); measurement reportability used the quality-score tiers above. All tasks, outputs, and thresholds are tabulated in Supplementary Table S3.

### Differences between development and evaluation data

Segmentation and EF were evaluated on held-out clips drawn from the same datasets (within-distribution), except for MIMIC-IV segmentation, which serves as an external generalisation cohort. Every evaluation clip was removed from training by identifier; patient-level separation is a stronger requirement, and we audited it per cohort rather than assuming it (Supplementary Note 2). The audit found that 27 of the 249 RVENet patients contributing RVEF evaluation clips also supplied training clips — in every case different loops of the same echocardiogram — so the RVEF evaluation is clip-disjoint rather than patient-disjoint, and Results reports a sensitivity analysis excluding those patients. The audit also found an identifier-format mismatch through which 26 CAMUS clips whose frames appear in training had escaped exclusion; those clips were removed and the CAMUS and pooled Dice re-scored under normalised identifiers (Table 2). Subject mapping in MIMIC-IV was completed using the dataset’s record list: all 686 held-out clips resolve to a subject, 41 of whom also contributed training clips, and a sensitivity analysis excluding them changes no chamber’s Dice by more than 0.001. Both audits are quantified in Supplementary Note 2. The RVEF reference (RVENet) contributed to training, so RVEF evaluation is same-centre, and no cross-centre RV test set was available. External validation therefore differs by task: segmentation was validated externally on MIMIC-IV; PH prediction was developed and evaluated entirely within MIMIC-IV; RVEF has no external cohort. Eligibility, outcome definitions, and inputs were otherwise identical between development and evaluation.

**Table 2.**
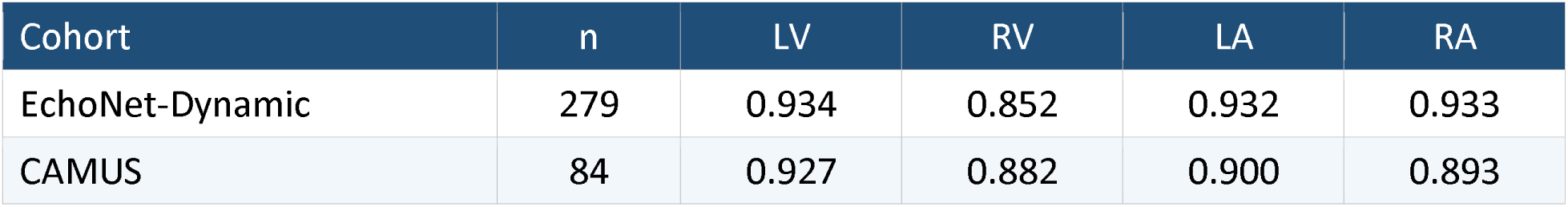
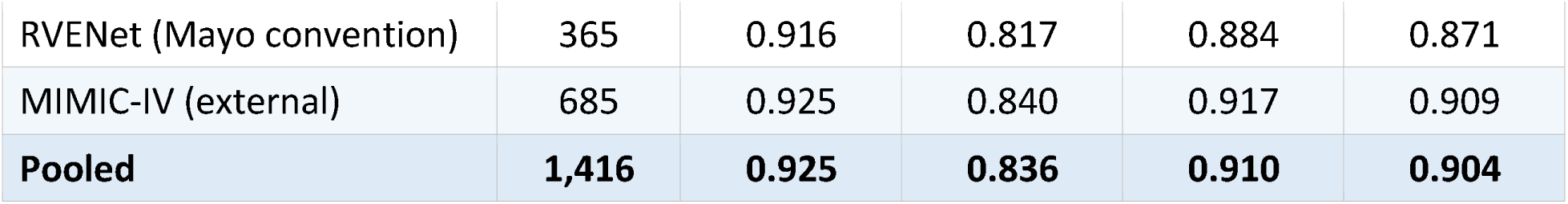
Four-chamber Dice by dataset and chamber (held-out, training-excluded; n=1,416). Mean Dice; pooled patient-clustered 95% CIs: LV 0.925 (0.922–0.927), RV 0.836 (0.830–0.841), LA 0.910 (0.906–0.914), RA 0.904 (0.899–0.908); per-cohort intervals and per-chamber n in Supplementary Table S8, clustering detail in Note 3. The per-cohort n counts clips with an LV mask; the pooled row counts the 1,416 clips with any annotated chamber, so the column does not sum. CAMUS RV/RA use the investigator-annotated subset (n=60).

### Model architecture and training

PH-ECHO-AI (“phechoVF”) is a single network with one shared encoder feeding four task heads (Fig. 1). A Swin Transformer V2-Small encoder processes 16-frame A4C clips (224×224); a norm-first temporal transformer integrates the cardiac cycle; and a feature-pyramid decoder with cohort-embedding modulation and domain-adaptive batch normalisation produces dense outputs. The heads emit five-class segmentation, seven landmark heatmaps (apices, septal and lateral annuli, septum), clip-level z-scored LVEF and RVEF, and auxiliary geometric features. Training used 8,416 clips (16,832 end-diastolic/end-systolic frames) under a staged curriculum with an EMA checkpoint and bfloat16 precision on a single GPU. Strain is computed from endocardial-contour wall paths (six LV segments; three RV free-wall segments) using a common set of valid edges at end-diastole and each frame, then averaged (segment-mean); LV strain is reported end-systolic and RV strain peak-systolic, per current guidance. This is Lagrangian contour-length strain from the segmented endocardial boundary: not speckle-tracking, and not validated against vendor strain per patient. We present it as a reproducible, vendor-independent deformation index, not a substitute for speckle-tracking strain (Limitations). TAPSE and MAPSE are obtained by projecting the tracked lateral annulus onto the end-diastolic long axis (in millimetres when DICOM calibration is present). Full inference and audit-correction detail appear in Supplementary Note 1. The RVEF head warrants explicit description because a two-dimensional A4C view supports fractional area change, not a volumetric ejection fraction. The head computes no geometric volume: it is a supervised regressor, trained on clips paired with 3D-echocardiographic RVEF labels from RVENet, that learns the mapping from four-chamber appearance and motion to the volumetric reference and applies it to new clips at inference. It therefore infers, rather than measures, the outflow-tract contribution, and it inherits the accuracy ceiling of its reference standard.

**Figure 1.**
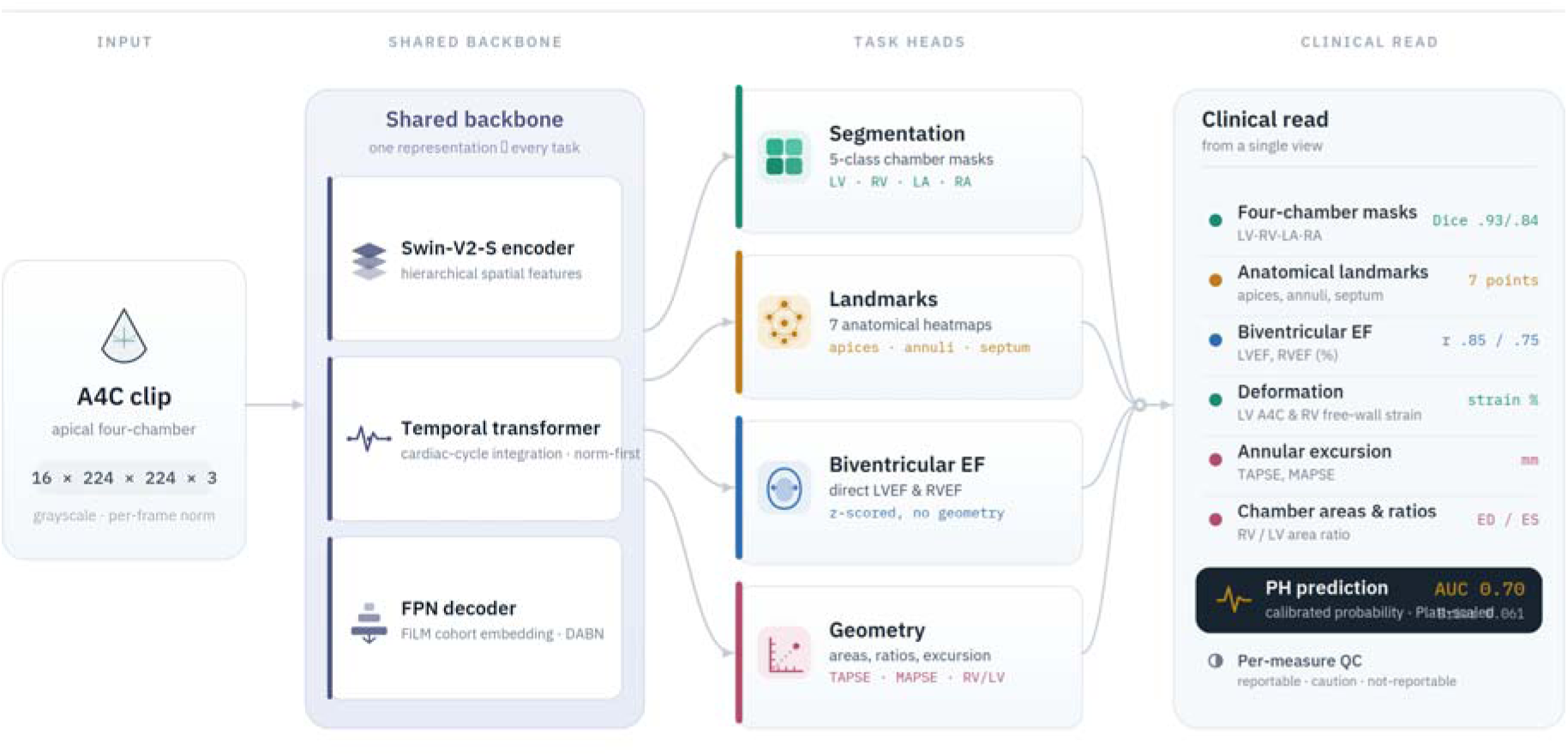
PH-ECHO-AI pipeline. One shared encoder and temporal transformer feed four heads (segmentation, landmarks, biventricular EF, geometry). A single A4C clip yields chamber masks, anatomical landmarks, LVEF/RVEF, deformation (LV A4C longitudinal strain, RV free-wall strain), annular excursion (TAPSE, MAPSE), chamber areas/ratios, and per-measure quality control.

## Ethics approval

The study uses de-identified, publicly released or credentialed-access datasets under their respective data-use agreements and operates under institutional research ethics approval (University Health Network / CAPCR #26-5021). A waiver of individual informed consent applies to the retrospective analysis of de-identified data, consistent with the originating datasets’ governance.

## Results

### Participants and flow

After excluding training clips by identifier and applying quality control, the held-out evaluation comprised 1,416 segmentation clips (EchoNet 279, CAMUS 84, RVENet 368, MIMIC 685; counting clips with any annotated chamber), 600 function/deformation clips (250 EchoNet, 350 RVENet), and a PH cohort of 1,718 patients of whom 1,076 entered the primary analysis (72 confirmed PH, 1,004 controls; 642 suspected-PH patients were excluded to keep the label objective; composition in Supplementary Table S6). The RVEF reference population spans elite athletes and healthy volunteers through heart failure and valvular disease (Supplementary Table S7). Counts per analysis accompany each result.

### Model development and specification

The released artefacts comprise the trained model weights and the complete training, inference, and evaluation code, which together permit prediction in new individuals and independent third-party evaluation; the model is released (not proprietary) under the terms stated in Open science. Participants and events per analysis were: segmentation n=1,416; LVEF n=250; RVEF n=350; PH n=1,076 (72 events); WHO Group 2 versus Group 3 classification, n=67 — the confirmed-PH subset with unambiguous WHO group assignment; a within-PH subtyping analysis, not a second detection cohort.

### Four-chamber segmentation performance

On 1,416 held-out, training-excluded clips, segmentation was accurate and maintained across cohorts it was not optimised for, including the Mayo-convention RVENet videos and the external MIMIC clinical clips (Table 2; Figs. 2–3; per-cohort CIs in Supplementary Table S8). Pooled Dice was 0.925 (LV), 0.836 (RV), 0.910 (LA), and 0.904 (RA); LV matched the 0.92 of EchoNet-Dynamic while extending delineation to three further chambers, and the RV, as expected the hardest, reached 0.836. Orientation robustness was mechanistic: 59% of RVENet clips are stored with the RV on the image right (RVENet is 98.6% Mayo by acquisition) and every CAMUS clip is stored in a non-canonical orientation, yet pooled stored-versus-canonical Dice differed by less than 0.001, with no RVENet or MIMIC chamber changing by more than 0.003.

**Figure 2.**
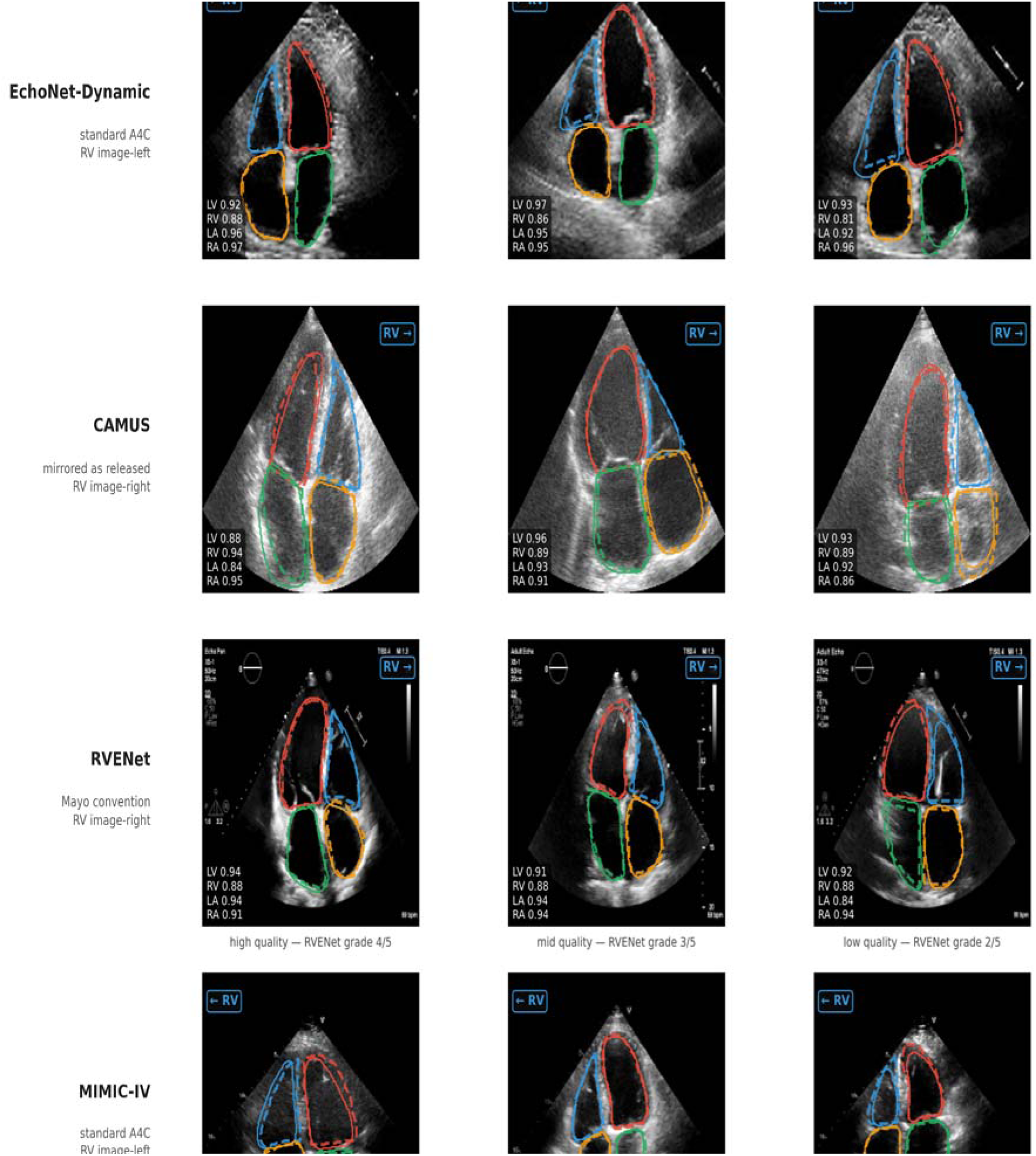
Representative randomly chosen segmentations across all four datasets. Predicted contours versus expert ground truth (three examples per cohort), illustrating consistent delineation across acquisition conventions and image quality, including the inverted Mayo-convention RVENet geometry and noisier MIMIC clinical clips. Panels appear in stored acquisition orientation, each with an orientation glyph derived from its own ground-truth mask centroids; every RVENet panel is stored Mayo (RV on the image right; RVENet is 98.6% Mayo by acquisition), and the row spans high-, mid-and low-quality acquisitions. Panels were sampled at random under a fixed seed, so several are deliberately poor — predicted and expert contours remain closely superimposed on exactly these clips. Per-panel Dice is inset.

**Figure 3.**
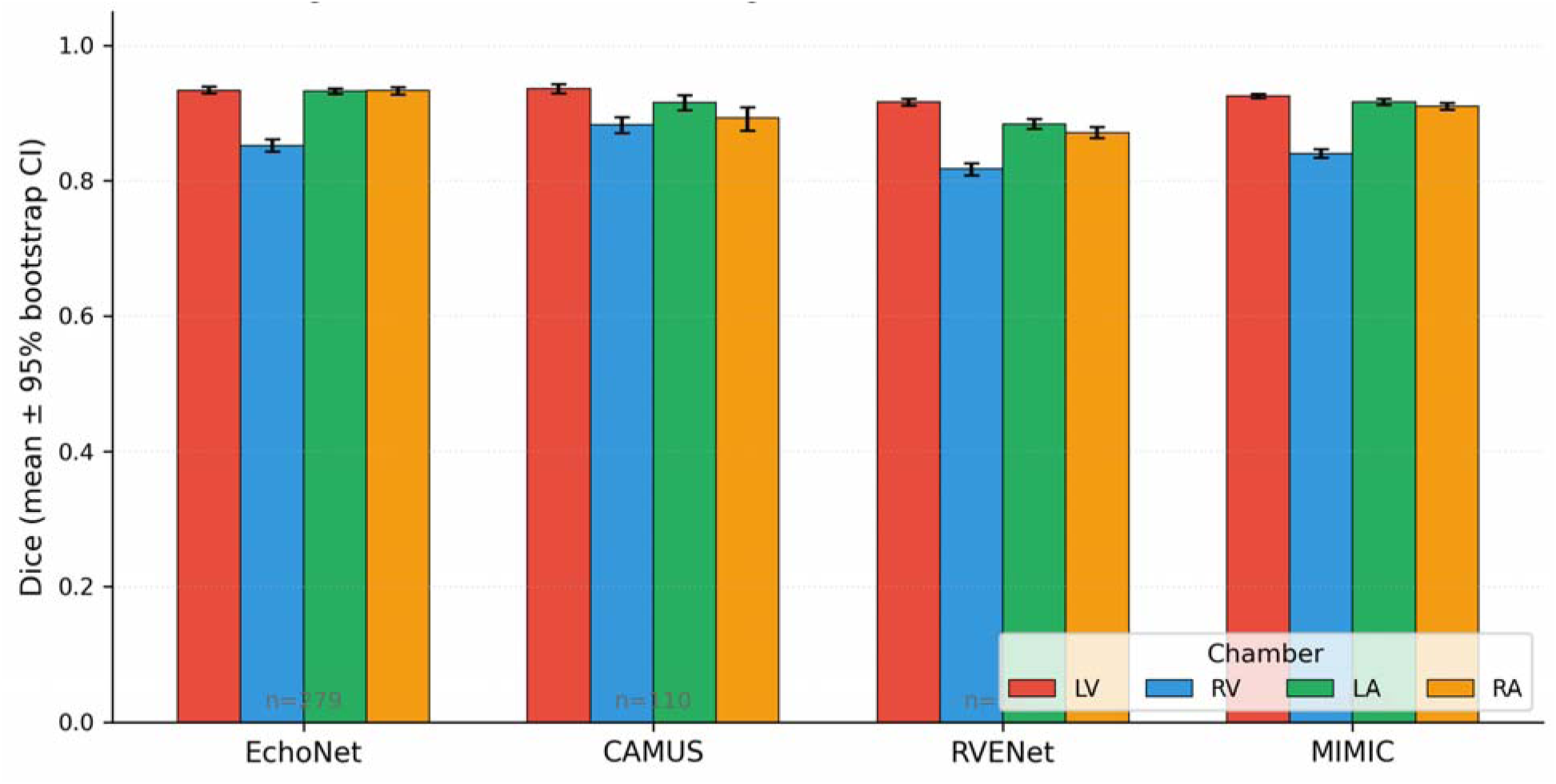
Per-cohort, per-chamber Dice with 95% bootstrap confidence intervals. Accuracy is high and tightly bounded across all four chambers and datasets; the right-sided chambers (RV, RA) are systematically hardest but remain well above the threshold required for reliable downstream measurement.

### Biventricular ejection-fraction performance

LVEF was estimated with r=0.845, MAE 4.67%, and minimal bias (−1.52%; n=250); RVEF was estimated directly against 3D echocardiography with r=0.754, MAE 4.98%, and bias −1.37% (n=350) (Table 3; Fig. 4). LVEF R² was 0.714 (95% CI 0.618 to 0.785) and RVEF R² 0.569 (95% CI 0.477 to 0.650). The RVEF intervals contain the R²=0.500 and MAE 5.06% of the dedicated single-network model of Tokodi et al. on the same reference standard, so the two cannot be separated statistically, whereas both LVEF intervals exclude the EchoNet-Dynamic values (R²=0.81; MAE 4.1%), so the residual gap to the LV-specialised network is real. Limits of agreement span roughly 25 ejection-fraction points for each ventricle, so agreement for an individual study is far looser than the MAE conveys; these outputs suit screening and cohort description, not serial tracking within one patient. On identical clips within identical resamples, the learned RVEF head exceeded RVFAC by 0.475 in correlation and 11.30 MAE points (both p<0.001; RVFAC vs 3D RVEF r=0.278, MAE 16.28%), and the learned LVEF head exceeded area-length estimation by 0.126 in correlation (p<0.001; Supplementary Note 3). RVEF accuracy was stable across acquisition-quality grade (r 0.74–0.89; MAE 3.45–5.35%) and across standard versus RV-focused A4C views (r 0.75 vs 0.76) (Supplementary Table S5). A prespecified sensitivity analysis excluding the 27 training-exposed patients (317 clips from 222 patients) left the correlation unchanged at 0.754 (95% CI 0.689 to 0.809) and raised MAE to 5.11% (95% CI 4.67 to 5.61); of the benchmarking comparisons, only the ensemble value of Tokodi et al. (4.57%) is sensitive to the restriction (Supplementary Note 2).

**Figure 4.**
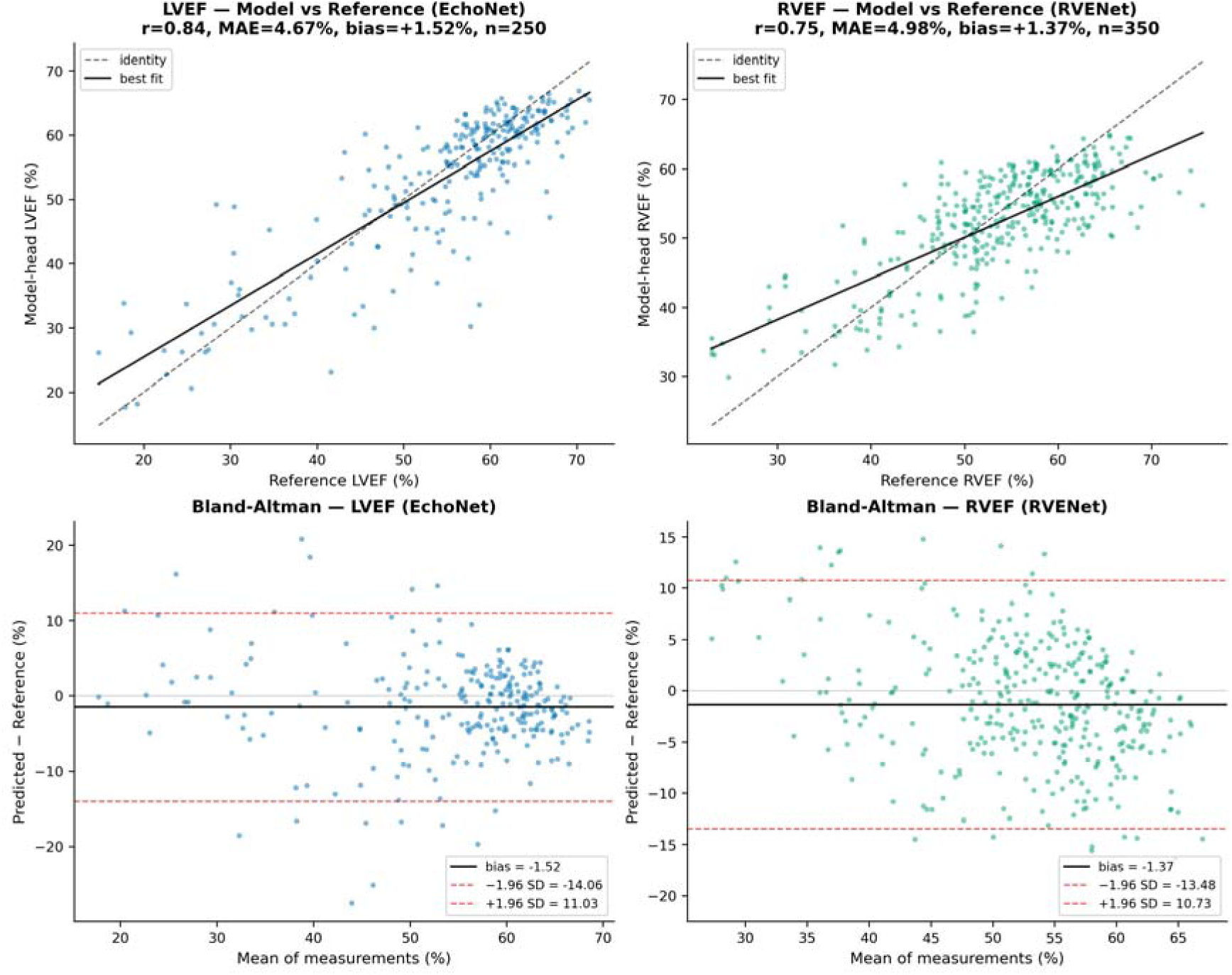
Biventricular EF validation. Scatter (with identity line) and Bland–Altman agreement for LVEF (EchoNet reference) and RVEF (RVENet 3D-echo reference). Both heads are accurate, near-unbiased, and well-distributed; RVEF agreement is achieved directly against the 3D gold standard available in these datasets; CMR, the definitive reference for right-ventricular volumes, was unavailable and anchors the planned external validation.

**Table 3.** Biventricular ejection-fraction validation (held-out). Direct model heads versus geometric baselines. Intervals are patient-clustered BCa bootstrap (Supplementary Note 3). LoA, 95% limits of agreement, spanning roughly 25 EF points per ventricle. The EchoNet-Dynamic reference LVEF is single-plane method of disks, the only view that dataset releases. The area-length row is a geometric baseline, not a clinical comparator. Subgroup performance in Supplementary Table S5.

| Measure (reference) | n | Pearson r | Spearman $\rho$ | MAE (%) | Bias (%) | 95% LoA (%) |
| --- | --- | --- | --- | --- | --- | --- |
| **LVEF, model head (EchoNet)** | 250 | 0.845 (0.786 to 0.886) | 0.787 | 4.67 (4.16 to 5.30) | -1.52 | -14.1 to +11.0 |
| LVEF, area-length (EchoNet) | 250 | 0.719 (0.576 to 0.795) | 0.692 | 9.37 (8.51 to 10.40) | +7.69 | -10.6 to +26.0 |
| **RVEF, model head (RVENet 3D)** | 350 | **0.754 (0.690 to 0.806)** | 0.688 | 4.98 (4.54 to 5.45) | -1.37 | -13.5 to +10.7 |
| RVFAC, geometric (RVENet 3D) | 350 | 0.278 (0.159 to 0.409) | 0.270 | 16.28 (15.19 to 17.50) | -12.14 | -42.0 to +17.7 |

### Benchmarking against published echocardiographic AI

Positioned against the principal published systems (Table 4), the unified model’s LVEF MAE (4.67%, 95% CI 4.16 to 5.30; R²=0.714, 95% CI 0.618 to 0.785) sits within roughly half a percentage point of the LV-specialised EchoNet-Dynamic (MAE 4.1%, R²=0.81) and PanEcho (MAE 4.4%), despite dedicating one of four heads to LVEF; the gap is real but modest, the visible price of consolidation. For the right ventricle the picture reverses. RVEF against 3D echocardiography is statistically indistinguishable from the dedicated single-network model of Tokodi et al. on the same RVENet standard and, on the full evaluation set, approaches their three-network ensemble, while EchoNet-RV reports the prognostically weaker RVFAC surrogate, which on our masks tracks 3D RVEF only modestly (r=0.278). Further dedicated RV models are in preparation; these comparisons describe the published record at the time of writing.

**Table 4.** Head-to-head comparison with published echocardiographic AI. Numbers as reported by each source on its respective test data; RVENet 3D-echo RVEF is the shared right-heart reference. R² for PH-ECHO-AI is r², with patient-clustered bootstrap intervals (Supplementary Note 3). The table reflects the published record at the time of writing. n/r, not reported / not applicable.

| System (year) | Scope | LV function | RV function |
| --- | --- | --- | --- |
| **PH-ECHO-AI** | Unified multi-task (4-chamber segmentation + landmarks + biventricular EF + deformation + PH) | LVEF $r=0.845$, MAE 4.67%, 95% CI 4.16 to 5.30; $R^2=0.714$ (95% CI 0.618 to 0.785) | RVEF $r=0.754$, MAE 4.98% (95% CI 4.54 to 5.45), $R^2=0.569$ (95% CI 0.477 to 0.650); direct regression against 3D echocardiography |
| EchoNet-Dynamic, Ouyang et al. (2020) | LV only (seg + LVEF) | LVEF MAE 4.1%, $R^2=0.81$; LV Dice 0.92 | n/r |
| RVEF-from-2D, Tokodi et al. (2023) | RV only (RVEF) | n/r | RVEF $R^2=0.500$ (single net), MAE 5.06% / 4.57% (ensemble); RVEF<45% acc 78% |
| EchoNet-RV, Tokodi/Ouyang et al. (2026) | RV only (seg + RVFAC) | n/r | RVFAC<35% AUC 0.86 / 0.73 / 0.68 (int./ext.); > EchoPrime, PanEcho |
| PanEcho, Holste et al. (2025) | View-agnostic foundation (39 tasks) | LVEF MAE 4.4% | RV indices (RVIDd MAE 4.0 mm); categorical RV function |

### Deformation and annular excursion

From the same masks and landmarks, the model computed vendor-independent endocardial-contour deformation and annular excursion that were physiologically coherent (Supplementary Table S9; Supplementary Figs. S1–S3). RV free-wall strain showed the expected strong inverse relationship with RVFAC (r=−0.640) and consistent inverse associations with reference RVEF (r=−0.392) and TAPSE (r=−0.418 in the 346 clips carrying pixel calibration); LV strain correlated with reference LVEF (r=−0.530). Landmark-derived excursion tracked ventricular function on both sides, more strongly on the right than on the left (TAPSE vs reference 3D RVEF r=0.530, n=346; MAPSE vs reference LVEF r=0.341, n=249). The asymmetry is expected rather than anomalous: EchoNet-Dynamic, which supplies every LVEF reference, is released without pixel calibration, so MAPSE on those clips is an uncalibrated pixel proxy rather than a millimetre measurement, and the LV correlation is attenuated accordingly. Strain timing was handled per guidance: end-systolic for the LV and peak-systolic for the RV. This asymmetry was empirically vindicated: for the RV, switching from peak to end-systolic timing halved the correlation with reference RVEF (r=−0.39 to −0.17), whereas the LV was timing-insensitive (Supplementary Fig. S2).

### Self-reported quality control

Each measurement carries a quality score (Supplementary Fig. S4; Supplementary Table S2). On curated reference data the model is essentially always reportable (LV and RV quality median 1.00); on unselected MIMIC clinical clips the same metrics fall and spread (RV quality median 0.82, 5th centile 0.49), discriminating interpretable A4C anatomy from off-axis and non-A4C acquisitions. This internal signal provides a practical, model-native mechanism for handling poor-quality or non-A4C inputs at deployment.

### Pulmonary-hypertension prediction performance

Confirmed PH is the clinically meaningful outcome and is reported in full in Table 5 (Fig. 5). Using echocardiographic geometry alone, with no clinical variables and no Doppler, RVSP-or catheter-confirmed PH (72 events / 1,076 patients) was detected with AUC 0.697 (95% CI 0.629 to 0.754) at sensitivity 0.47 and specificity 0.84. Discrimination was lower for the noisier composite any-PH label (AUC 0.595, 95% CI 0.568 to 0.622; n=1,718), consistent with medical-population “controls” harbouring undiagnosed PH. Gradient boosting (AUC 0.645) could not be separated from logistic regression at 72 events (difference 0.052, 95% CI −0.011 to +0.120, p=0.12). Platt recalibration preserved discrimination (AUC 0.682) and cut the Brier score from 0.214 to 0.061, yielding interpretable absolute risks. Permutation analysis ranked LV A4C strain, RVFAC, RV free-wall strain, RVEF, and TAPSE as most informative (Supplementary Fig. S5; Supplementary Table S10). Within confirmed PH, World Health Organization (WHO) Group 2 versus Group 3 was separable (accuracy 0.88, 95% CI 0.79 to 0.94; macro-F1 0.63, 95% CI 0.47 to 0.87; n=67), although Groups 1, 4, and 5 were too rare to model (Supplementary Fig. S6). These analyses establish feasibility rather than a deployable PH classifier.

**Figure 5.**
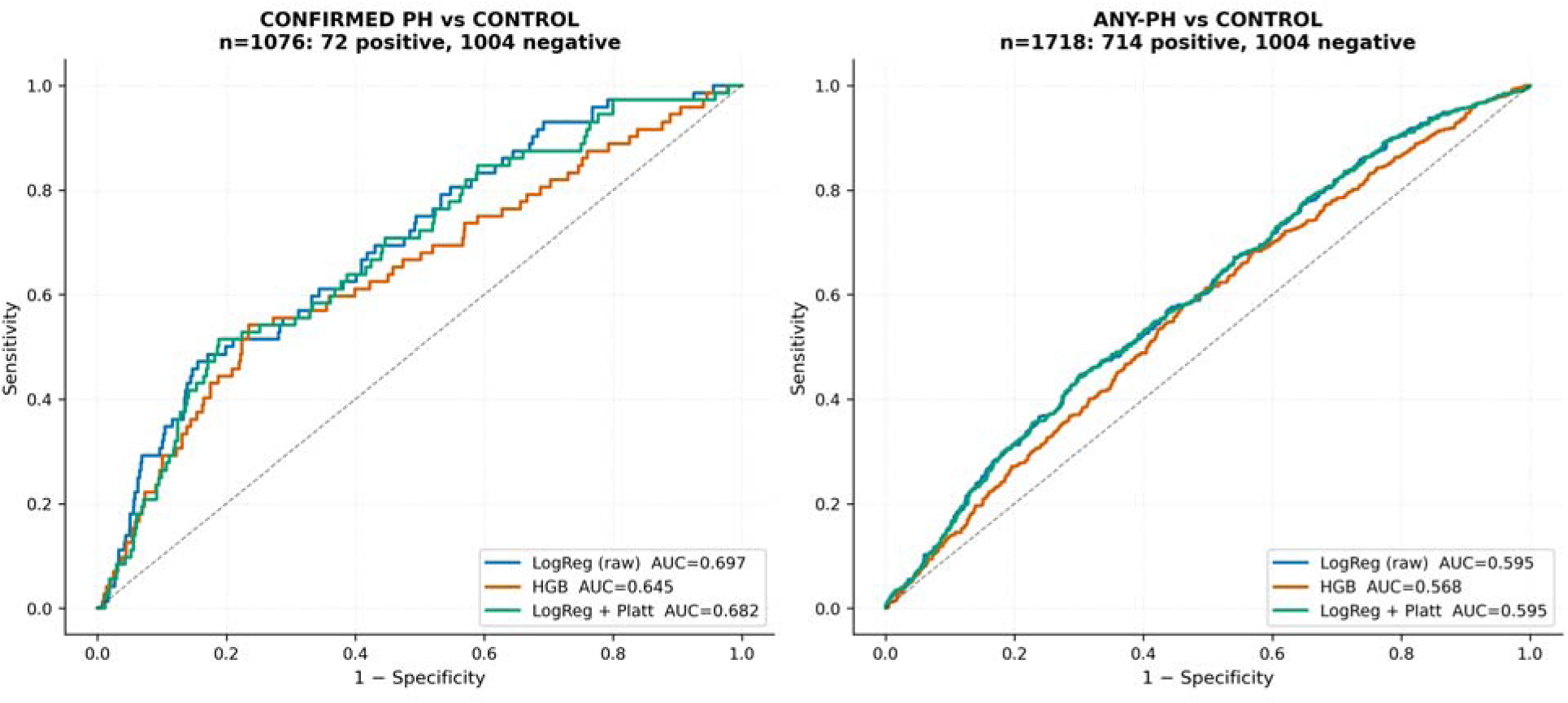
PH-prediction ROC curves. Receiver-operating-characteristic curves for confirmed-PH and any-PH detection across three classifiers using echocardiographic geometry alone; discrimination is markedly stronger for haemodynamically confirmed PH. Probability calibration and feature importance are shown in Supplementary Figures S5 and S7.

**Table 5.** Pulmonary-hypertension prediction performance (main analysis). Echocardiographic geometry alone; no clinical variables, no Doppler. Confirmed PH required PH support plus an objective threshold (RVSP ≥40 mmHg or catheter PASP ≥35 mmHg). Sensitivity and specificity are at the cross-validated Youden point; intervals are patient-level BCa bootstrap with the Youden threshold re-derived per resample (Supplementary Note 3), and DeLong intervals agree within 0.006. Platt recalibration is a one-parameter sigmoid fitted within nested cross-validation. Gradient boosting and logistic regression cannot be separated (ΔAUC 0.052, 95% CI −0.011 to +0.120, p=0.12); the gradient-boosting AUC varies with scikit-learn version (0.645 under the analysis environment, 0.642 under the pinned release environment). The final row is within-PH subtyping in the 67 patients with unambiguous WHO group, so accuracy and macro-F1 replace sensitivity and Brier. Calibration and importance in Supplementary Figures S5 and S7 and Table S10. n/r, not reported.

| Analysis | Outcome | n (events) | AUC (95% CI) | Operating point (sens. / spec.) | Brier score (95% CI) |
| --- | --- | --- | --- | --- | --- |
| Logistic regression (primary) | RVSP- or catheter-confirmed PH | 1,076 (72) | 0.697 (0.629 to 0.754) | 0.47 (0.32–0.57) / 0.84 (0.50–0.91) | 0.214 (0.203 to 0.225) |
| Logistic regression, Platt-recalibrated | RVSP- or catheter-confirmed PH | 1,076 (72) | 0.682 (0.612 to 0.741) | 0.51 (0.37–0.75) / 0.81 (0.41–0.86) | 0.061 (0.050 to 0.074) |
| Gradient boosting | RVSP- or catheter-confirmed PH | 1,076 (72) | 0.645 (0.568 to 0.711) | 0.54 (0.39–0.67) / 0.77 (0.61–0.82) | 0.071 (0.057 to 0.087) |
| Logistic regression | Any-PH composite label | 1,718 (714) | 0.595 (0.568 to 0.622) | 0.45 (0.25–0.60) / 0.70 (0.34–0.85) | 0.243 (0.239 to 0.248) |
| Within-PH subtyping | WHO Group 2 vs Group 3 | 67 | n/r | accuracy 0.88 (0.79–0.94) | macro-F1 0.63 (0.47–0.87) |

### Model updating (recalibration)

Platt recalibration constituted the only model updating: raw class-balanced probabilities were over-confident, and a one-parameter sigmoid restored calibration to the true event rate while preserving discrimination (Supplementary Fig. S7).

## Discussion

We designed and implemented a single, fully reproducible model that performs comprehensive right ventricle-focused echocardiographic interpretation from one standard A4C view. Our model segments all four chambers, estimates both ventricular ejection fractions directly, computes physiologically coherent deformation and annular excursion, and supports PH prediction, without requiring multiple models (4, 6, 9, 14).

RV systolic function is the strongest echocardiographic predictor of survival in PH and across cardiopulmonary disease (1), and among RV indices, 3D-echocardiographic RVEF carries the greatest prognostic weight. In a meta-analysis of 1,928 patients, each one–standard-deviation reduction in 3D-RVEF was associated with a 2.6-fold increase in adverse events and was a significantly stronger predictor than TAPSE, FAC, or free-wall strain in head-to-head comparison (16); in pulmonary arterial hypertension specifically, 3D-RVEF correlates with invasive hemodynamics more closely than any other non-invasive index, independently predicts clinical events, and identifies markedly worse survival (15).

RVFAC, a measurement provided by a commercial automated RV tool (9, 14), is a 2D surrogate whose agreement with volumetric RVEF is view-dependent and only moderate (apical four-chamber RVFAC versus CMR-derived RVEF r≈0.50) and does not account for the right-ventricular outflow-tract contribution to ejection, potentially leading to errors in estimates of global RV function (17, 18). Automation amplifies these limitations: a fully automated workflow recently reported an 11–14% RVFAC bias even as other measurements remained accurate (19). Our own single-view automated RVFAC behaved similarly (r=0.278 against 3D RVEF), whereas the learned RVEF head reached r=0.754 (R²≈0.57). A model that estimates RVEF directly therefore returns the more clinically meaningful, and more reliably automatable, quantity. One qualification is owed: our model cannot image the outflow tract either. Unlike RVFAC, which discards the outflow-tract contribution by construction, the head is trained against a reference that includes it and so infers it — statistically, not anatomically. The inference should hold where inflow and outflow covary, as in the acquired disease that dominates training, and fail where they dissociate: congenital disease, after right-ventricular outflow-tract surgery, arrhythmogenic cardiomyopathy. Validation in those populations remains outstanding.

### Relationship to prior automated echocardiography

Automated echocardiography of the left ventricle has matured rapidly. For example, EchoNet-Dynamic achieved cardiologist-level LVEF estimation (4). Furthermore, a subsequent blinded, randomised trial found that initial AI assessments were not only non-inferior but superior to expert sonographers, as cardiologists were less likely to revise the AI reads. This demonstrates that well-validated echo AI is ready for prospective clinical testing (5). Multi-task foundation models broadened the task list but comparatively provide fewer metrics of the right ventricle (6, 7). For the right ventricle specifically, our RVEF accuracy (R²=0.569, 95% CI 0.477 to 0.650) is statistically indistinguishable from the dedicated single-network model of Tokodi et al. on the identical RVENet 3D standard (R²=0.500) and, on the full evaluation set, approaches their three-network ensemble (9), whereas EchoNet-RV, the current state of the art for RV segmentation, optimises the prognostically weaker RVFAC surrogate (8). To our knowledge, PH-ECHO-AI is the first model to deliver four-chamber segmentation, anatomical landmarks, both ventricular ejection fractions, deformation, excursion, and PH prediction from a single A4C clip in one forward pass.

### Deformation and excursion

RV free-wall longitudinal strain is among the most reproducible and prognostically powerful RV indices, outperforming TAPSE, FAC, and estimated pulmonary pressure for predicting mortality across PH aetiologies (3). A free-wall strain worse (less negative) than −20% predicts death even when TAPSE is preserved (20), and forms the basis for our −20% reporting threshold. Our endocardial-contour, vendor-independent measurement of strain reproduced these expected relationships (free-wall strain versus RVFAC r=−0.640; versus reference RVEF r=−0.392) and recovered annular excursion (TAPSE versus 3D RVEF r=0.530) despite the known angle dependence of TAPSE (20–22). Because vendor speckle-tracking strain remains incompletely standardised across machines (21, 23), a reproducible mask-derived alternative is attractive. It is important to emphasise, however, that contour strain is mathematically distinct from speckle-tracking and is not numerically interchangeable (24–26).

### Pulmonary-hypertension detection

Echocardiographic PH screening conventionally rests on the tricuspid-regurgitation jet velocity (TRV), and existing automated tools follow suit. An automated TRV pipeline reached 73% accuracy against catheterisation in the ASPIRE cohort (27), and a fully automated deep-learning echo workflow detected PH with AUC 0.75–0.79 (19), performance bounded by the fact that echo-estimated and invasive pressures agree in fewer than 60% of cases (28). Against this backdrop, detecting catheter-or RVSP-confirmed PH with an AUC of approximately 0.70 from chamber geometry and function alone is a meaningful result. Deliberately excluding the TRV signal is consistent with the broader literature applying AI to PH across the electrocardiogram and chest radiograph (28, 29). We frame PH prediction as proof of concept (72 events) and anticipate that adding Doppler TRV and clinical variables would materially raise discrimination. The clinical case for a geometry-only route does not rest on outperforming Doppler: a measurable TRV is absent in a large minority of studies. Among 1,262 patients studied within two days of right-heart catheterization, 36% had no reported TRV, and 47% of those had invasively confirmed PH (31); pulmonary artery acceleration time, estimated mean pressure and arterial diameter are likewise often unavailable. A geometry-only read therefore serves the patients the conventional Doppler pathway silently fails.

### Generalisation and the multi-task, domain-adaptive design

Echocardiography models are highly sensitive to domain shifts, and naïvely merging datasets risks shortcut learning (30). To address this, we trained across four datasets and two acquisition conventions using cohort-conditioned normalisation and domain-adaptive batch statistics. This approach yielded robust segmentation on external MIMIC and RVENet clips, showing at most a 0.003 Dice change between orientations. While these results validate our architectural choices, geographically external right ventricular validation remains necessary and is addressed next. One design limitation deserves note: every measurement here is contour-based, derived from an explicit endocardial segmentation. That buys auditability — each number traces to a contour a clinician can inspect and overrule — and inherits the segmentation’s ceiling. Self-supervised latent-predictive architectures such as EchoJEPA, pretrained on 18 million unlabelled studies, learn without drawing a boundary and may in time exceed contour-derived measurement (32); we regard the two as complementary, the contour route retaining an interpretability latent representation do not yet offer.

### Limitations

RVEF was validated on training-excluded RVENet clips, but RVENet contributed to training and no cross-centre RV test set was available. The evaluation is clip-disjoint rather than patient-disjoint: 27 of 249 contributing patients supplied clips to both sides, always from the same study, so the model had seen a different cardiac cycle of the recording it was scored on. Excluding them changes the correlation by 0.000 and the MAE by 0.13 points, so the effect is small, but leakage should be assumed absent only where audited; the CAMUS identifier mismatch (repaired, with the affected clips excluded and re-scored) and the MIMIC-IV subject mapping (completed: 41 of 686 evaluation subjects overlap training, shifting no chamber by more than 0.001) are documented in Supplementary Note 2. The reference standard was 3D echocardiography rather than CMR, so our accuracy is bounded by 3D echocardiography itself and agreement with CMR is untested. Given the well-documented internal-to-external performance drop in echocardiographic AI (8, 30) (the dedicated RVEF model showed RVEF R² 0.500 to 0.329 from internal to external testing (9)), an analogous decline should be anticipated and prospectively quantified.

LVEF and RVEF outputs are highly correlated (r≈0.97), reflecting genuine physiological covariation but raising the possibility of shared signal; RVEF is nonetheless validated independently against 3D-echo RVEF. Consistent with prior catheter analyses in this programme, the model captures systolic function, not pulmonary pressure, and does not replace invasive haemodynamics.

Strain is mask-derived and mathematically distinct from vendor speckle-tracking: values are internally coherent but not interchangeable, no per-patient deformation ground truth exists, and the prognostic value established for vendor strain cannot be assumed to transfer. Our thresholds are borrowed from that literature as a starting point; the outcome studies needed to confirm equivalence have not been done.

End-diastole and end-systole are defined by extremes of LV mask area rather than valve-closure events; absolute millimetre excursion depends on available pixel calibration (scale-free for EchoNet).

With 72 events across eleven features, the geometry-only PH model (confirmed-PH AUC ∼0.70) is proof of concept, not a deployable tool; WHO Groups 1, 4, and 5 and severity were too rare to model, and A4C selection in MIMIC used a model-native quality filter rather than a dedicated validated view classifier.

### Usability in current care and next steps

At deployment, poor-quality or non-A4C inputs are flagged by the model’s per-measure quality tiers, which mark non-interpretable acquisitions as not reportable rather than returning spurious values. The system requires minimal user interaction, loading an A4C clip and, where available, entering DICOM pixel calibration, and is designed for use by non-expert and point-of-care operators with clinical oversight; it is intended to augment, not replace, expert interpretation. Priority next steps are (i) cross-centre external validation of RVEF against cardiac magnetic resonance, the definitive reference standard for right-ventricular volumes, and against 3D echocardiography, for which an institutional cohort of approximately 800 studies with paired CMR-derived RVEF has been identified; (ii) prospective evaluation against clinical outcomes; (iii) a multi-annotator inter-observer study; (iv) a complete sociodemographic fairness evaluation; and (v) a dedicated validated A4C view classifier to replace the quality-filter surrogate.

## Conclusion

A single unified deep-learning model delivers comprehensive right-heart-focused echocardiographic interpretation from one apical four-chamber view: four-chamber segmentation, direct biventricular ejection fraction competitive with specialised models, vendor-independent deformation and excursion, and feasible pulmonary-hypertension prediction, reproducibly and across multiple datasets and acquisition conventions. By targeting RVEF directly and consolidating the right-heart read into one model and one view, PH-ECHO-AI provides a practical foundation for scalable RV screening, with prospective cross-centre validation as the priority next step.

### Reporting and supplementary material

This study is reported in accordance with TRIPOD+AI (Collins et al., BMJ 2024) and CLAIM 2024. The Supplementary Material contains the completed TRIPOD+AI checklists (Appendices 1 and 2), Supplementary Tables S1–S10, Supplementary Figures S1–S7, and Supplementary Notes 1–3 covering inference and audit corrections, the patient-level separation audit, and interval estimation.

### Open science

Supported in part by the pulmonary-hypertension research programme at Toronto General Hospital / University Health Network; funders had no role in study design, analysis, or reporting. The authors declare no competing interests. No prospective protocol was prepared; a pre-specified analysis plan and code are released. The study was not registered (retrospective development/evaluation using existing data). EchoNet-Dynamic, CAMUS, and RVENet are available from their providers under their licences; MIMIC-IV-ECHO is available to credentialed PhysioNet users. All analysis code (preprocessing, training, evaluation) is openly released with a pinned environment; because every source dataset prohibits redistribution of individual-level derivatives, the derived annotations and trained model weights are distributed through a credentialed deposit rather than from the corresponding author. The released code reproduces every reported value under a fixed random seed in the pinned environment.

### Patient and public involvement

There was no formal patient or public involvement in the design, conduct, or reporting of this methodological development study. Patient and public engagement is planned for the prospective clinical-validation phase.

## Data Availability

Code and model will be made available upon publication.

