## Supplement for "Development and multi-dataset evaluation of a unified single-view deep-learning model for the right heart: four-chamber segmentation, biventricular ejection fraction, deformation, and pulmonary-hypertension prediction from the apical four-chamber echocardiogram"

### Supplementary Note 1. Inference scheme

Two-pass inference. For each clip, the whole-video left-ventricular (LV) mask-area curve (Gaussian-smoothed) identifies the strongest-contraction beat (end-diastole → end-systole → end-diastole). Ejection fraction is then computed from a single canonical 16-frame full-cycle forward pass, which is the regime in which the EF heads were trained; per-frame segmentation and landmarks are separately aggregated across dense, 50%-overlapping 16-frame windows (up to 32 frames per beat). This separation is deliberate: naïvely averaging EF over partial-cycle windows degraded LVEF MAE from 4.7 to 8.3 percentage points and bias from −1.5 to −7.0, whereas the two-pass design fully restored EF performance. Landmark coordinates are obtained by spatial soft-argmax with a confidence estimate; the right-ventricular (RV) mask undergoes morphological cleaning; inference uses bfloat16 mixed precision.

Audit corrections to the deformation pipeline. The deformation and quality-control code underwent a structured audit; the following nine corrections were implemented and are reflected in all reported values:

1. Nomenclature: “LV GLS” was replaced by “LV A4C longitudinal strain” throughout, because only the apical four-chamber plane is sampled (not a true global longitudinal strain).
2. Asymmetric timing: the displayed LV strain is end-systolic (ASE/EACVI guidance) and the displayed RV free-wall strain is peak-systolic (RV consensus document); this asymmetry was empirically vindicated (Fig. S2; main text).
3. Segment-mean global strain: per-segment strain curves are computed and then averaged, replacing a length-weighted whole-polyline formula that could be biased by a single noisy segment.
4. Endpoint re-pinning (a): anatomical anchors (mitral/tricuspid annulus and apex landmarks) are re-pinned to model-predicted coordinates after temporal smoothing.
5. Endpoint re-pinning (b): because the RV free-wall path is short, a one-pixel endpoint drift materially changes wall length; re-pinning after smoothing removes this artefact.
6. Same valid edges across frames: strain at end-diastole and at each frame is computed over the intersection of valid contour edges, eliminating a failure mode in which dropping different edges at the two time points falsely appeared as shortening.
7. Dense overlapping windows: up to 32 frames per beat with 50%-overlap 16-frame windows feed per-frame segmentation and landmarks, while EF is taken from the separate canonical full-cycle pass (above).
8. Real model-mask quality control: a quality score combining mask-boundary distance, frame-to-frame length jump, endpoint jump, segment survival, and landmark confidence (with reportable/caution/not-reportable tiers) replaced a placeholder that always returned the maximum score.
9. Provenance: deformation outputs are explicitly labelled as endocardial-contour strain (not speckle-tracking) in the output metadata.

Robustness of the headline measurements to these corrections is quantified in Table S4: the EF heads were unchanged, the RV free-wall-strain–versus–RVFAC relationship strengthened, and the LV strain–versus–LVEF relationship changed only marginally—the expected price of switching to the more rigorous, guideline-concordant computation.

### Supplementary Note 2. Patient-level separation audit

Every evaluation clip was removed from training by clip identifier, and no evaluation clip appears in the training manifest in any cohort. Patient-level separation is a stronger requirement, and we audited it per cohort rather than assuming it. This note reports the audit in full; the main text summarises it in Methods (Differences between development and evaluation data) and Limitations.

RVENet (RVEF evaluation). 27 of the 249 patients contributing the 350 evaluation clips also supplied training clips, and in all 27 cases the training and evaluation clips come from the same study: different loops of a single echocardiogram sharing one volumetric label. The RVEF evaluation is therefore clip-disjoint rather than patient-disjoint. Two alternative explanations were tested and rejected: no evaluation identifier matches any training identifier under normalisation (excluding an identifier-format artefact), and no PatientHash is null, empty, or mixes sexes (excluding a codebook collision). A sensitivity analysis excluding all 27 patients leaves 317 clips from 222 patients: correlation is unchanged at 0.754 (95% CI 0.689 to 0.809) and mean absolute error rises from 4.98% to 5.11% (95% CI 4.67 to 5.61), both far inside the confidence intervals. One benchmarking comparison is sensitive to the restriction: the 4.57% ensemble value of Tokodi et al. lies inside the full-set MAE interval but outside the clean-subset interval, whereas their 5.06% single-network value lies inside both. A leakage-exploitation test found no coherent advantage on exposed patients: across the four chambers the exposed-minus-clean Dice differences do not agree in sign, and the right ventricle moves in the direction opposite to what memorisation would produce.

CAMUS (segmentation evaluation). 50 of the 151 ground-truth files carry an _ED or _ES suffix while the training manifest stores the unsuffixed clip name, and the original exclusion filter matched exact strings, so 26 held-out clips whose unsuffixed identifier is a training clip — the same annotated frames the model trained on — were not excluded. Those clips scored above the clean CAMUS remainder (LV +0.039, LA +0.063; Mann-Whitney p<10⁻⁹), the expected signature of memorisation, and they have been removed. Every evaluation entry point now excludes training clips through a shared canonical identifier that strips the phase suffix, an automated check refuses to score any set containing a training identifier, and a regression test asserts that the 26 known-bad identifiers collide with their training spellings. The re-scored CAMUS cohort comprises 84 clips from 65 patients: LV 0.927 (95% CI 0.918 to 0.934) and LA 0.900 (0.885 to 0.911); the investigator-annotated RV/RA subset (n=60) contained none of the affected clips and is unchanged. Pooled Dice moves by less than 0.001 in every chamber (LA 0.911 to 0.910 at three decimals; LV, RV, RA unchanged).

MIMIC-IV (segmentation evaluation). Filenames carry a study identifier but no subject. The MIMIC-IV-ECHO record list distributed with the dataset resolves all 686 held-out clips to a subject; the derived feature table used in the original audit reached only 231, and the two routes agree on every shared clip. Of the 686 evaluation subjects (each contributing one evaluation clip), 41 also contributed training clips, and 37 of those evaluation clips come from the same study as a training clip. A sensitivity analysis excluding all 41 clips changes no chamber’s Dice by more than 0.001 (largest shift +0.0006, RA). The previously reported lower bound of 10 is superseded by this complete measurement.

EchoNet-Dynamic. The dataset releases no patient identifier; the one-clip-per-patient structure is assumed by the processing pipeline rather than established by the data.

### Supplementary Note 3. Interval estimation and paired comparisons

All confidence intervals reported in the manuscript are bias-corrected and accelerated (BCa) bootstrap intervals from 10,000 resamples under seed 20260804, resampling contributing patients rather than clips; the acceleration term uses a delete-one-patient jackknife. Clips from one patient share a reference label, so their errors are correlated and a clip-level interval is too narrow. This matters materially for RVENet, whose 350 function clips derive from 249 patients (168 with one clip, 64 with two, 14 with three, 3 with four) and whose 365 segmentation clips derive from 99 patients; EchoNet-Dynamic contributes one clip per patient, so its intervals are unaffected by the choice. Patient-clustered intervals are wider than the Fisher z intervals they replaced in every case, and both benchmarking conclusions survive the widening.

Sensitivity and specificity intervals for the pulmonary-hypertension model re-derive the Youden threshold within each resample, so they carry threshold-selection uncertainty and are correspondingly wide. DeLong intervals agree with the bootstrap on every AUC to within 0.006.

Paired comparisons evaluate both members of a pair on identical clips within identical resamples, so the interval is on the difference itself. The learned RVEF head exceeds RVFAC by 0.475 in Pearson correlation (95% CI 0.370 to 0.584) and undercuts it by 11.30 MAE percentage points (95% CI 10.17 to 12.48), both p<0.001, on the identical 350 clips. The learned LVEF head exceeds the area-length estimate by 0.126 in correlation (95% CI 0.059 to 0.269; p<0.001) on the identical 250 clips. Against fixed published values, our LVEF MAE interval (4.16 to 5.30) excludes the 4.1% of EchoNet-Dynamic, whereas our RVEF MAE interval (4.54 to 5.45) contains both the 5.06% single-network and 4.57% ensemble values of Tokodi et al. Logistic regression and gradient boosting differ by 0.052 in AUC (95% CI −0.011 to +0.120, p=0.12) on identical resamples and cannot be separated at 72 events.

### Supplementary Tables

**Table S1. Data sources and study design.** Four single-centre cohorts spanning two acquisition conventions and four centres across three countries (TRIPOD+AI items 5a–6b, 16). Prediction tasks, outcomes, and thresholds are detailed in Supplementary Table S3.

| Dataset (centre, country) | Reported dates | Role | Held-out clips | Reference standard |
| --- | --- | --- | --- | --- |
| EchoNet-Dynamic (Stanford, USA) | 2016–2018 | LV reference + LV training | 279 (Dice) / 250 (function) | Reference LVEF (single-plane method of disks); expert LV tracings |
| CAMUS (St-Étienne, France) | Not stated | 4-chamber segmentation GT | 84 (Dice) | Expert LV/LA + investigator 4-chamber masks |
| RVENet (Semmelweis, Hungary) | 2013–2021 | RVEF reference + 4-chamber GT | 365 (Dice) / 350 (function) | 3D-echocardiography RVEF; expert masks |
| MIMIC-IV-ECHO (Beth Israel, USA) | MIMIC-IV window | External generalisation + PH cohort | 685 (Dice) / 1,076 (PH) | Expert masks; ICD/RVSP/catheter PH labels |

**Table S2. Model-mask quality-control distribution by dataset.**

Landmark confidence and per-chamber quality scores on curated reference data versus unselected MIMIC clinical clips, showing the score discriminates interpretable A4C anatomy (cited in Methods, data preparation; and Results, self-reported quality control).

| Dataset | Metric | Median | Q05 | Q25 | Q75 | Q95 |
| --- | --- | --- | --- | --- | --- | --- |
| *MIMIC clinical (n=5,270)* | Landmark confidence | 368 | 102 | 219 | 645 | 1,076 |
| *MIMIC clinical (n=5,270)* | LV quality | 0.87 | 0.44 | 0.72 | 0.97 | 1.00 |
| *MIMIC clinical (n=5,270)* | RV quality | 0.82 | 0.49 | 0.68 | 0.94 | 1.00 |
| **Curated reference (n=600)** | Landmark confidence | 1,074 | 776 | 969 | 1,166 | 1,273 |
| **Curated reference (n=600)** | LV quality | 1.00 | 1.00 | 1.00 | 1.00 | 1.00 |
| **Curated reference (n=600)** | RV quality | 1.00 | 0.95 | 1.00 | 1.00 | 1.00 |

**Table S3. Prediction tasks, outcomes, reference standards, and decision thresholds.**

EF and PH outcomes are concurrent/diagnostic; no prognostic time horizon applies. Strain and excursion have no per-patient ground-truth label and were evaluated for physiologic coherence (cited in Methods, outcomes and reference standards; and model output and decision thresholds).

| **Task** | **Output** | **Reference standard** | **Threshold** |
| --- | --- | --- | --- |
| Four-chamber segmentation | Per-pixel class (LV/RV/LA/RA) | Expert-reviewed manual masks | — (Dice) |
| LVEF | Continuous % | Reference LVEF (EchoNet) | <50% dysfunction (sex-neutral; ASE/EACVI sex-specific limits 52% men, 54% women) |
| **RVEF (primary)** | Continuous % | 3D-echocardiography (RVENet) | <45% dysfunction |
| RVFAC | Continuous % | (vs 3D RVEF) | <35% dysfunction |
| LV / RV longitudinal strain | Continuous % | Physiologic coherence | LV worse (less negative) than −18%; RV worse (less negative) than −20% |
| TAPSE / MAPSE | Continuous mm | Physiologic coherence | Abnormal: TAPSE <17 mm; MAPSE <11 mm |
| RV/LV area ratio | Ratio | Physiologic coherence | ≥0.6 abnormal |
| Pulmonary hypertension | Calibrated probability | RVSP ≥40 (echo) or PASP ≥35 (cath) | Youden p=0.63 |

**Table S4. Robustness of headline measurements to the deformation audit.**

Pre-audit (initial pipeline) versus post-audit (corrected pipeline; Supplementary Note 1). Correlations are with the stated reference (cited in Methods, analytical methods).

| **Measure (correlation)** | **Pre-audit** | **Post-audit** | **Δ** |
| --- | --- | --- | --- |
| LVEF head vs reference LVEF (r) | 0.845 | 0.845 | 0.00 |
| RVEF head vs reference RVEF (r) | 0.754 | 0.754 | 0.00 |
| LV A4C strain vs reference LVEF (r) | −0.56 | −0.53 | +0.03 |
| RV free-wall strain vs reference RVEF (r) | −0.385 | −0.392 | −0.007 |
| RV free-wall strain vs RVFAC (r) | −0.562 | −0.640 | −0.078 |

**Table S5. RVEF accuracy by video-quality grade and A4C view type (RVENet function cohort).** Direct RVEF head versus 3D-echo reference, stratified (cited in Methods, Fairness; and Results, biventricular EF).

| **Stratum** | **n** | **Pearson r** | **Spearman ρ** | **MAE (%)** | **Bias (%)** |
| --- | --- | --- | --- | --- | --- |
| *Video quality 1 (lowest)* | 13 | 0.838 | 0.660 | 4.46 | −1.09 |
| *Video quality 2* | 103 | 0.742 | 0.595 | 5.35 | −1.66 |
| *Video quality 3* | 158 | 0.737 | 0.675 | 4.97 | −1.24 |
| *Video quality 4* | 66 | 0.781 | 0.798 | 4.76 | −1.21 |
| *Video quality 5 (highest)* | 10 | 0.894 | 0.894 | 3.45 | −2.01 |
| **Standard A4C view** | 254 | 0.753 | 0.701 | 4.84 | −1.08 |
| **RV-focused A4C view** | 96 | 0.761 | 0.658 | 5.35 | −2.14 |

**Table S6. Pulmonary-hypertension cohort composition and WHO-group breakdown (MIMIC-IV).**

Confirmed PH requires an objective threshold; controls have no PH support (cited in Results, Participants and flow).

| **Category** | **n** | **Definition** |
| --- | --- | --- |
| **Confirmed PH** | 72 | is-PH AND (echo RVSP ≥40 mmHg OR catheter PASP ≥35 mmHg) |
| Suspected PH | 642 | is-PH (ICD/text/medication) without an RVSP/PASP value |
| **Control** | 1,004 | No PH ICD code, text mention, or PH-specific medication |
| **WHO groups within confirmed PH** | — | Group 2 (n=60) · Group 3 (n=7) · Group 4 (n=3) · Group 1 (n=2) |

**WHO Group 2 versus Group 3 within confirmed PH: out-of-fold confusion matrix (n=67)**

| **True group** | **Predicted Group 2** | **Predicted Group 3** | **Total** |
| --- | --- | --- | --- |
| WHO Group 2 (left-heart disease) | 57 | 3 | 60 |
| WHO Group 3 (lung disease or hypoxia) | 5 | 2 | 7 |
| Total | 62 | 5 | 67 |

*Computed from the persisted out-of-fold predictions. Accuracy 0.881 (95% CI 0.791 to 0.940); macro-F1 0.634 (95% CI 0.468 to 0.867). The interval width is the finding: this analysis establishes feasibility and cannot support a claim about subtyping performance. Groups 1 and 4 (n=2 and n=3) were too rare to model, and Group 5 was absent (n=0).*

**Table S7. Reference RVEF by RVENet patient subgroup (function-evaluation cohort, n=350).** Participant characteristics across clinically meaningful subgroups (cited in Results, Participants and flow).

| **Subgroup** | **n** | **Reference RVEF, mean ± SD (%)** |
| --- | --- | --- |
| Healthy volunteers | 59 | 62.0 ± 4.3 |
| Paediatric healthy volunteers | 22 | 60.2 ± 5.3 |
| Athletes | 47 | 54.7 ± 6.5 |
| Non-compaction cardiomyopathy | 11 | 56.5 ± 6.2 |
| History of heart transplant | 25 | 52.2 ± 4.1 |
| Paediatric, history of kidney transplant | 9 | 60.1 ± 5.7 |
| Valvular heart disease (mitral) | 65 | 51.1 ± 6.4 |
| Valvular heart disease (aortic) | 41 | 49.5 ± 8.8 |
| Heart failure, reduced EF (HFrEF) | 33 | 37.0 ± 9.4 |
| Other | 38 | 56.4 ± 5.9 |

**Table S8. Per-cohort, per-chamber Dice with patient-clustered 95% bootstrap confidence intervals (held-out, training-excluded; n=1,416).**

CAMUS RV/RA evaluated on the investigator-annotated subset (n=60); the n column lists clips with an LV mask and, after the slash, the RV-annotated count where it differs (RVENet per-chamber n: LV 365, RV 366, LA 359, RA 360; 368 clips carry at least one annotated chamber); pooled excludes CAMUS RV/RA (cited in Results, four-chamber segmentation). Intervals are bias-corrected and accelerated, from 10,000 resamples of contributing patients under seed 20260804, so clips from one patient are resampled together. Clustering widens the RVENet intervals by 0.004 on average and the CAMUS intervals by 0.001–0.002; the other cohorts move by less than 0.001. RVENet and CAMUS contribute several clips per patient (365 clips from 99 patients; 84 clips from 65 patients, seven of them present under both an unsuffixed and an _ED/_ES spelling).

| Cohort | n | LV (95% CI) | RV (95% CI) | LA (95% CI) | RA (95% CI) |
| --- | --- | --- | --- | --- | --- |
| EchoNet-Dynamic | 279 | 0.934 [0.928–0.938] | 0.852 [0.842–0.860] | 0.932 [0.927–0.936] | 0.933 [0.926–0.938] |
| CAMUS | 84 / 60 | 0.927 [0.918–0.934] | 0.882 [0.868–0.893] | 0.900 [0.885–0.911] | 0.893 [0.864–0.905] |
| RVENet | 365 / 366 | 0.916 [0.909–0.922] | 0.817 [0.802–0.829] | 0.884 [0.874–0.891] | 0.871 [0.860–0.881] |
| MIMIC-IV | 685 | 0.925 [0.922–0.928] | 0.840 [0.832–0.846] | 0.917 [0.912–0.921] | 0.909 [0.904–0.914] |
| **Pooled** | **≤1,416** | **0.925 [0.922–0.927]** | **0.836 [0.830–0.841]** | **0.910 [0.906–0.914]** | **0.904 [0.899–0.908]** |

**Table S9. Deformation and annular-excursion coherence (held-out).**

Endocardial-contour strain and landmark-derived excursion against reference function and against one another (cited in Results, deformation and annular excursion).

| **Relationship** | **n** | **Pearson r** | **Spearman ρ** |
| --- | --- | --- | --- |
| RV free-wall strain (peak) vs RVFAC | 597 | −0.640 | −0.690 |
| RV free-wall strain (peak) vs reference RVEF | 350 | −0.392 | −0.382 |
| RV free-wall strain (peak) vs TAPSE (calibrated) | 346 | −0.418 | −0.440 |
| RV free-wall strain (peak) vs RV/LV area ratio | 598 | −0.169 | −0.199 |
| LV A4C strain (ES) vs reference LVEF | 250 | −0.530 | −0.541 |
| LV A4C strain (ES) vs MAPSE | 594 | −0.274 | −0.241 |
| TAPSE vs reference RVEF | 346 | 0.530 | n/r |
| MAPSE vs reference LVEF | 249 | 0.341 | 0.476 |

*MAPSE versus reference LVEF is attenuated relative to the corresponding TAPSE comparison because EchoNet-Dynamic, which supplies every LVEF reference, is released without pixel calibration, so MAPSE on those clips is an uncalibrated pixel proxy rather than a millimetre measurement. TAPSE comparisons use the 346 RVENet clips carrying DICOM calibration.*

**Table S10. Permutation feature importance for confirmed-PH prediction (gradient-boosted model, 10 repeats).**

Mean decrease in performance when each feature is permuted (cited in Results, PH prediction).

| **Rank** | **Feature** | **Mean importance** | **SD** |
| --- | --- | --- | --- |
| 1 | LV A4C longitudinal strain (ES) | 0.0147 | 0.0035 |
| 2 | RV fractional area change | 0.0109 | 0.0027 |
| 3 | RV free-wall strain (peak) | 0.0103 | 0.0023 |
| 4 | RVEF | 0.0066 | 0.0023 |
| 5 | TAPSE | 0.0066 | 0.0020 |
| 6 | LVEF | 0.0054 | 0.0020 |
| 7 | MAPSE | 0.0047 | 0.0014 |
| 8 | RV end-systolic area | 0.0025 | 0.0008 |
| 9 | LV end-diastolic area | 0.0019 | 0.0014 |
| 10–11 | RV end-diastolic area; RV/LV area ratio | 0.0000 | 0.0000 |

### Supplementary Figures

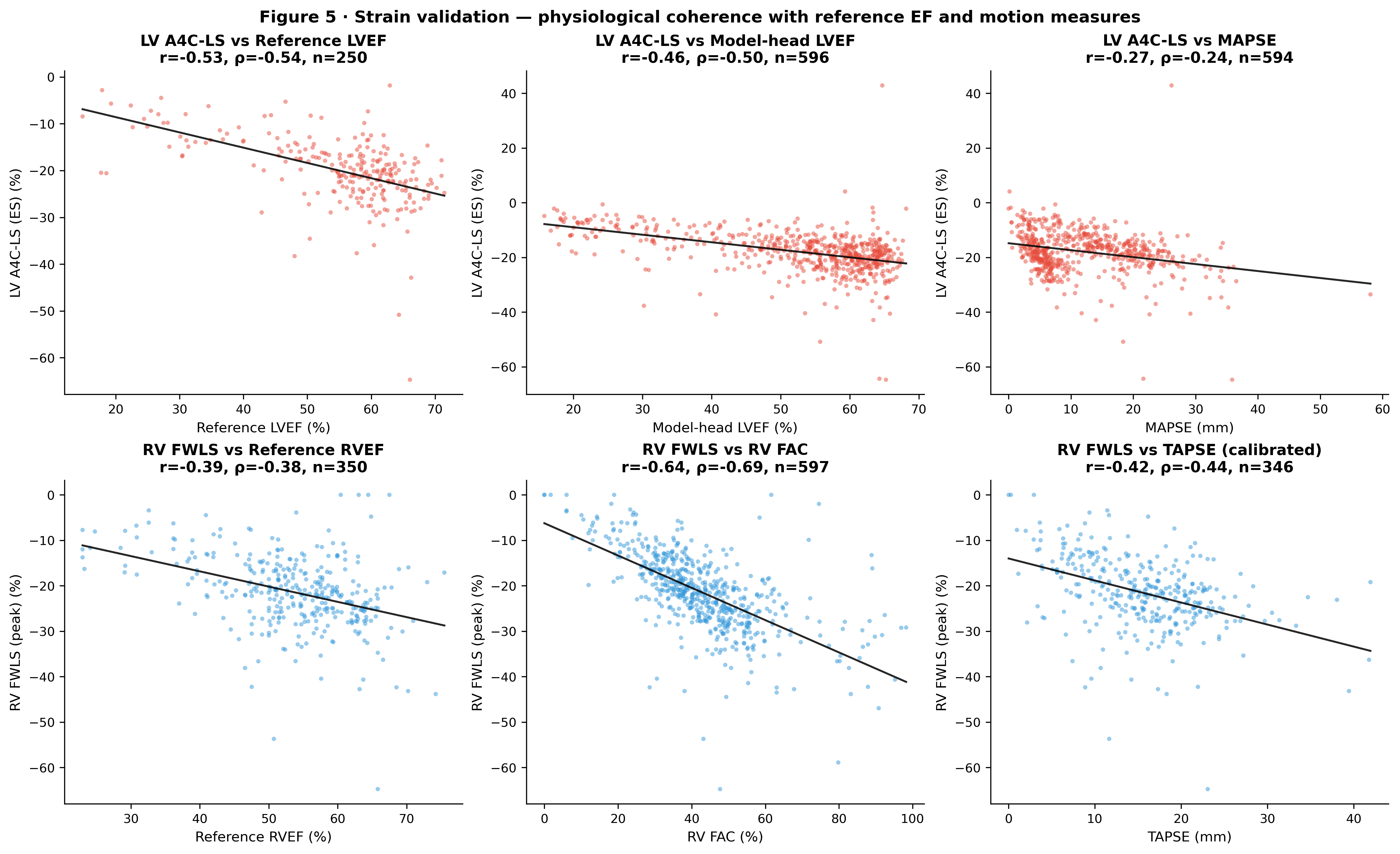

**Figure S1. Deformation validation.** Relationships between model-derived deformation/excursion indices and reference measures. RV free-wall strain tracks RVFAC and reference RVEF with the expected negative direction; LV strain tracks LVEF; landmark-derived TAPSE/MAPSE recapitulate ventricular function. (Cited in Results, deformation and annular excursion.)

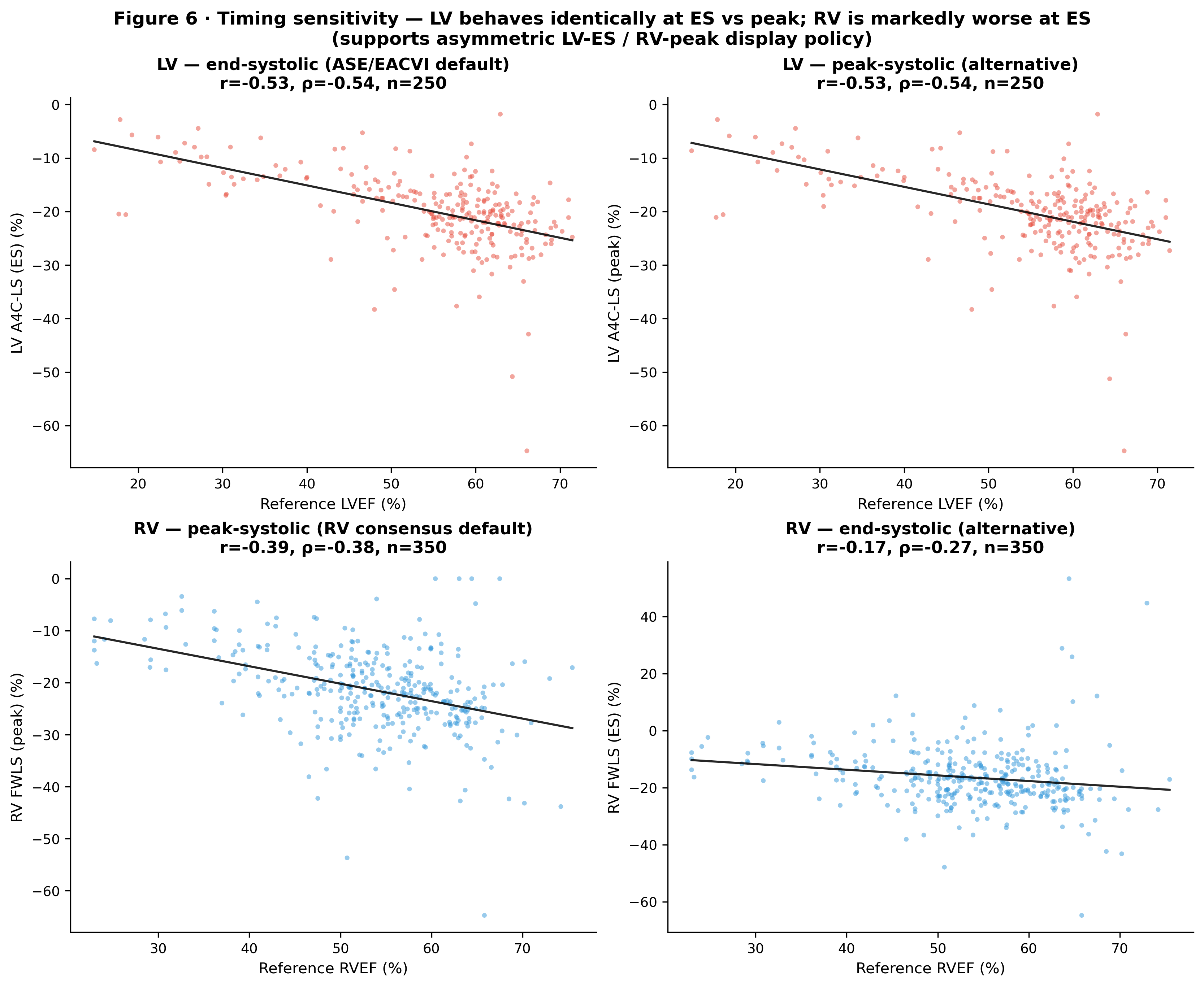

**Figure S2. Empirical validation of guideline-concordant strain timing.** LV strain is timing-insensitive, whereas RV strain correlates with reference RVEF roughly twice as strongly at peak-systolic than end-systolic timing, justifying the asymmetric reporting convention (LV end-systolic, RV peak-systolic). (Cited in Results, deformation and annular excursion.)

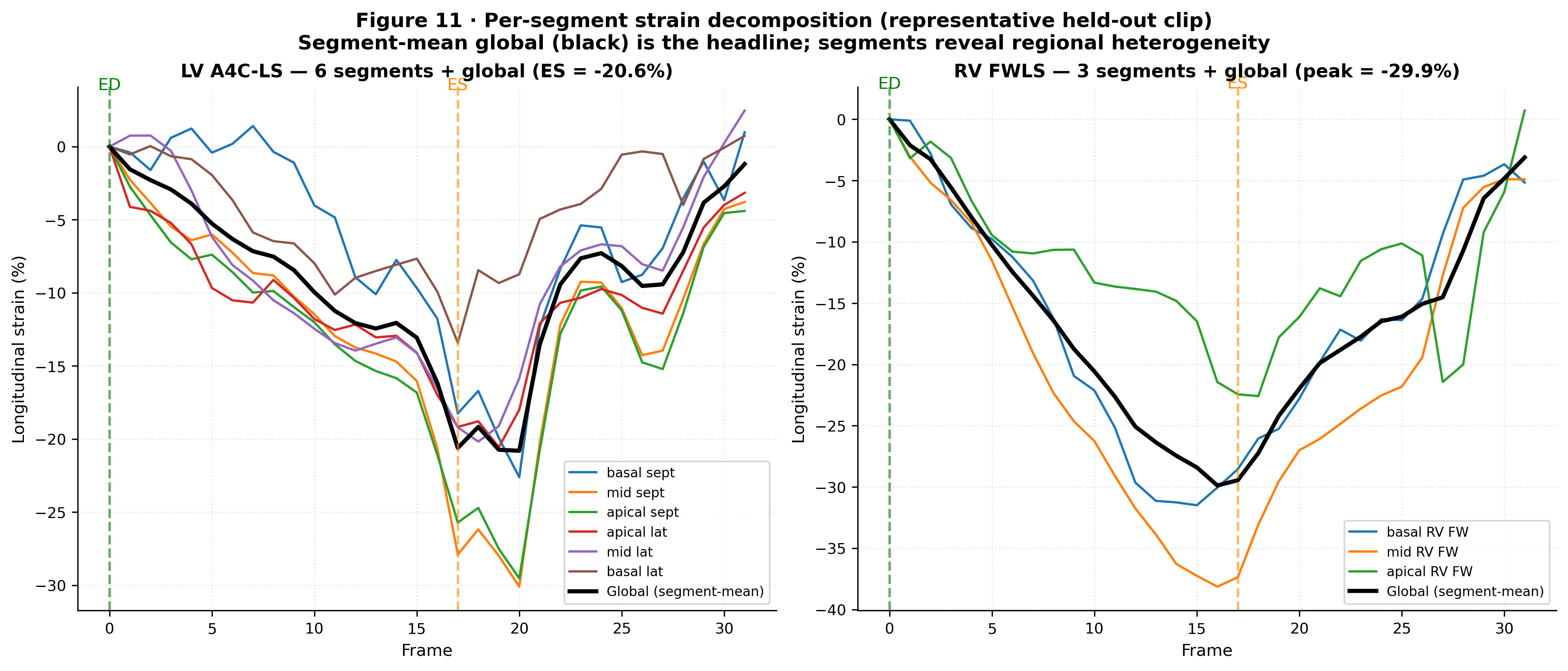

**Figure S3. Segmental deformation on a representative clip.** Six-segment LV and three-segment RV free-wall longitudinal strain curves and bullseye, illustrating the regional output of the deformation head. (Cited in Results, deformation and annular excursion.)

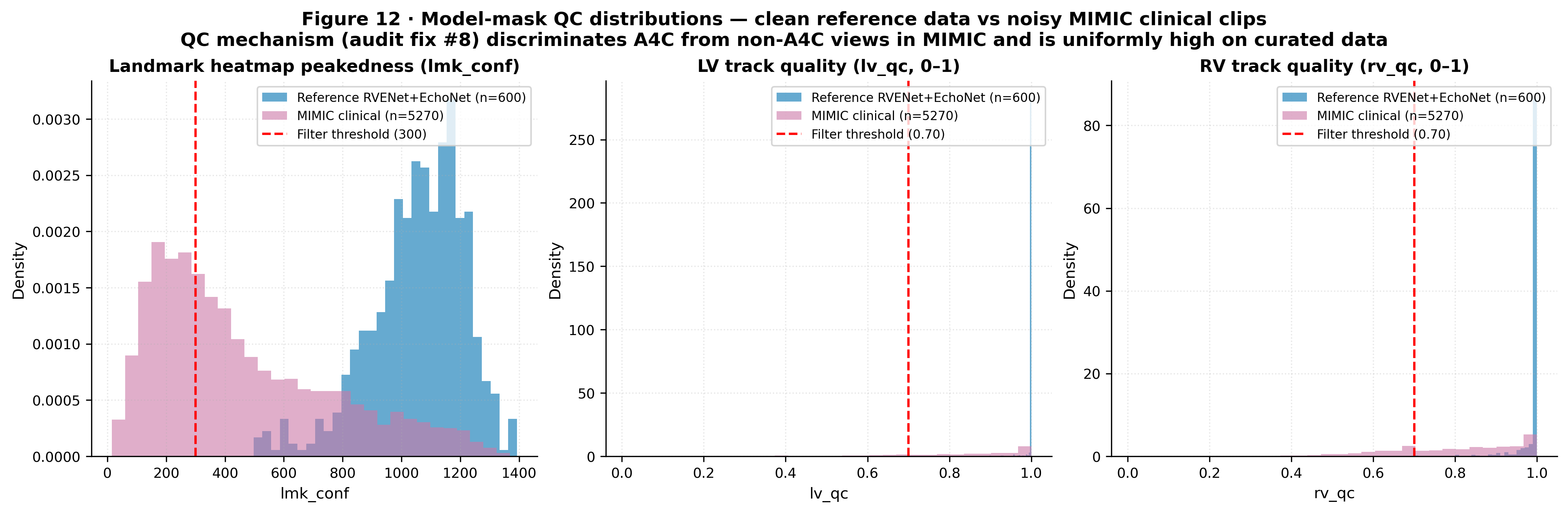

**Figure S4. Quality-control distributions.** Per-measure quality on curated reference data (near-ceiling) versus unselected MIMIC clinical clips (lower, wider), demonstrating that the model identifies non-interpretable acquisitions. (Cited in Results, self-reported quality control.)

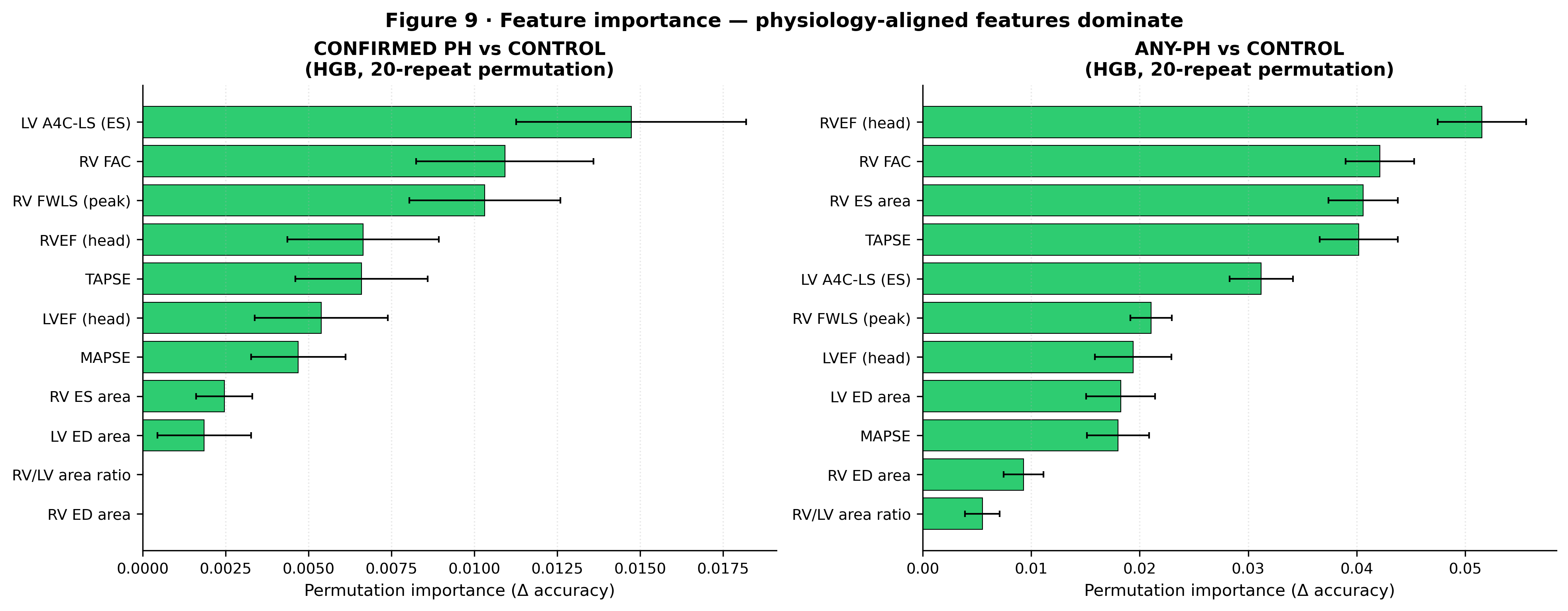

**Figure S5. Permutation feature importance for PH prediction.** A physiologically coherent signature dominated by ventricular-function and chamber-geometry features (LV A4C strain, RVFAC, RV free-wall strain, RVEF, TAPSE). (Cited in Results, PH prediction.)

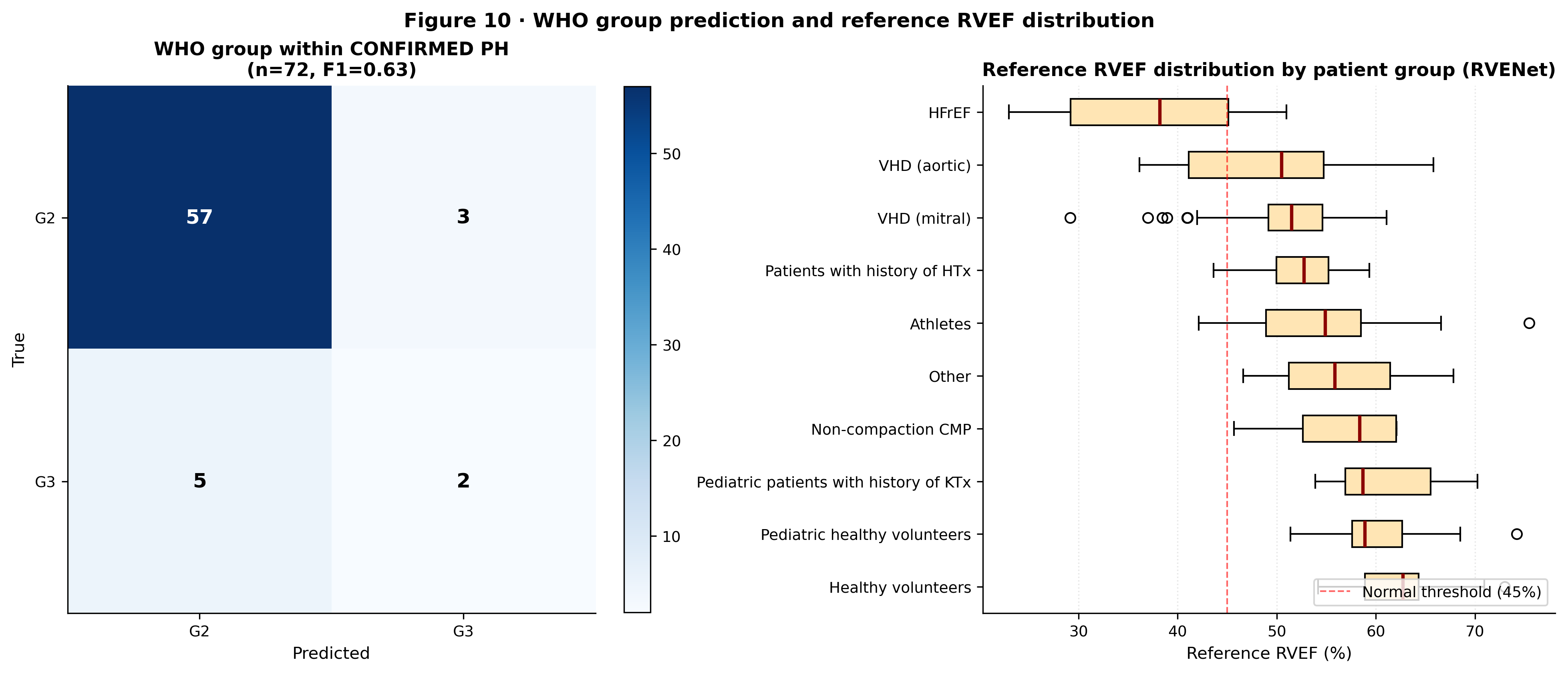

**Figure S6. WHO-group classification and reference distributions.** WHO Group 2 versus 3 confusion within confirmed PH, and reference RVEF by RVENet patient subgroup spanning athletes and healthy volunteers through heart failure and valvular disease. (Cited in Results, PH prediction and Participants.)

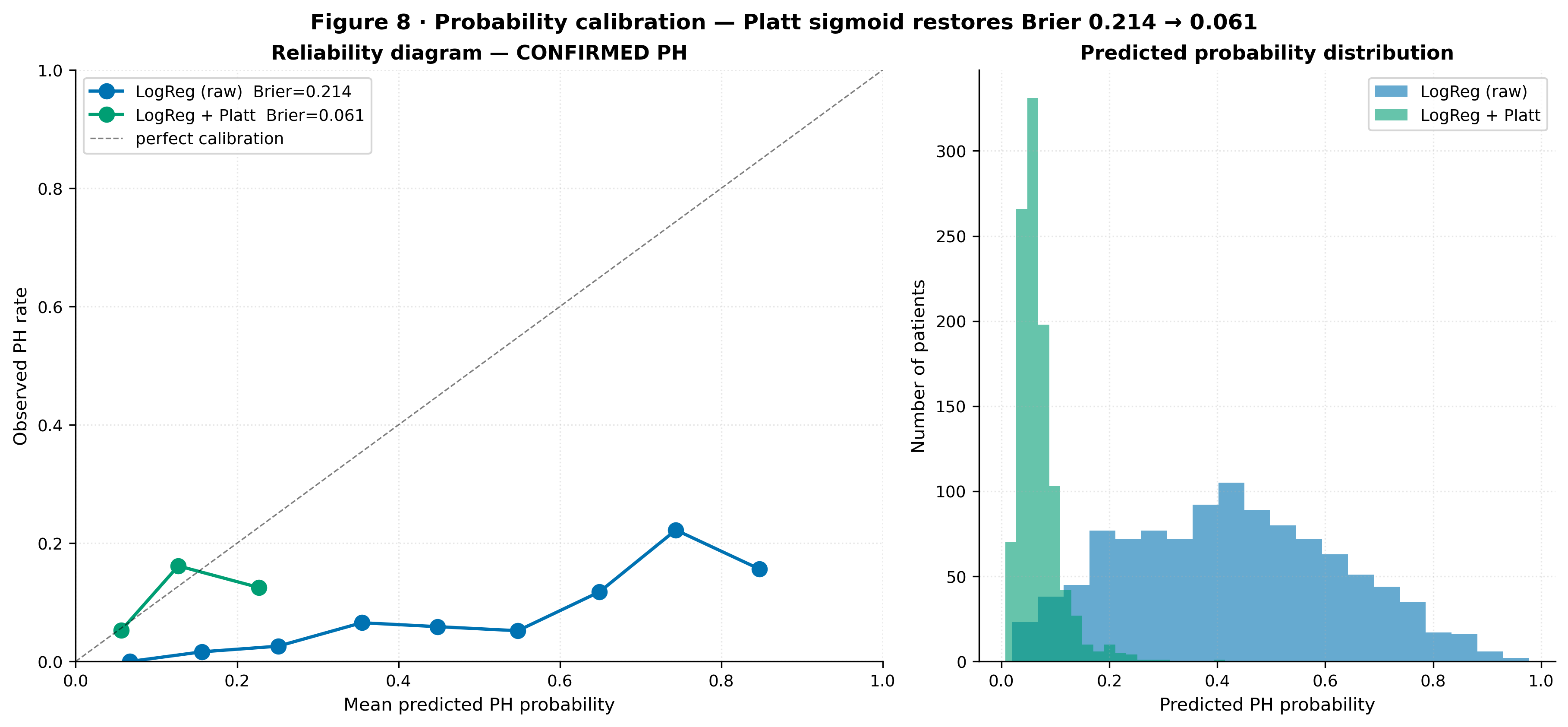

**Figure S7. Pulmonary-hypertension probability calibration.** Reliability diagram and predicted-probability histograms before and after Platt scaling. One-parameter sigmoid calibration restores agreement with the observed event rate (Brier 0.214 → 0.061) while preserving discrimination. (Cited in Results, PH prediction and model updating.)

### Appendix 1. Completed TRIPOD+AI checklist

*Item wording is abbreviated; full item text appears in Collins GS, et al. BMJ 2024;385:e078378. “Reported in manuscript” gives the section heading where each item is addressed; page numbers refer to the submitted manuscript. D, development; E, evaluation.*

| Item | Section / topic | Reported in manuscript |
| --- | --- | --- |
| 1 | Title | Title page (development + evaluation; target population; outcomes) — p. 1 |
| 2 | Abstract | Abstract (structured); see Appendix 2 — p. 2 |
| 3a | Background / rationale | Introduction — p. 4 |
| 3b | Target population / users | Introduction — p. 4 |
| 3c | Health inequalities | Introduction — p. 4 |
| 4 | Objectives | Introduction → Objectives — p. 4 |
| 5a | Data sources | Methods → Source of data and study design; Table S1 — pp. 4–5 |
| 5b | Participant dates | Methods → Source of data and study design; Table S1 — pp. 4–5 |
| 6a | Setting / centres | Methods → Participants, setting, and treatments; Table S1 — p. 5 |
| 6b | Eligibility criteria | Methods → Participants, setting, and treatments — p. 5 |
| 6c | Treatments | Methods → Participants (not applicable; cross-sectional) — p. 5 |
| 7 | Data preparation / QC | Methods → Data preparation and quality control — p. 5 |
| 8a | Outcome definition / timing | Methods → Outcomes and reference standards; Table S3 — pp. 5–6 |
| 8b | Outcome assessors | Methods → Outcomes and reference standards. Assessor demographic characteristics are not reported; single-annotator review is acknowledged in Limitations — pp. 5–6, 20 |
| 8c | Outcome blinding | Methods → Outcomes and reference standards — p. 6 |
| 9a | Predictor choice | Methods → Predictors / model inputs — p. 6 |
| 9b | Predictor definition / measurement | Methods → Predictors / model inputs — pp. 6, 8 |
| 9c | Predictor assessors | Methods → Predictors (no subjective interpretation at inference) — p. 6 |
| 10 | Sample size | Methods → Sample size — p. 6 |
| 11 | Missing data | Methods → Missing data — p. 6 |
| 12a | Data use / partitioning | Methods → Analytical methods — pp. 6–8 |
| 12b | Predictor handling | Methods → Analytical methods — p. 6 |
| 12c | Model type / building / internal validation | Methods → Analytical methods; Model architecture; Note 1 — pp. 6, 8 |
| 12d | Heterogeneity across clusters | Methods → Analytical methods (no cluster model fitted) — pp. 6, 10 |
| 12e | Performance measures | Methods → Analytical methods — p. 6 |
| 12f | Model updating | Methods → Analytical methods; Results → Model updating — pp. 6, 18 |
| 12g | Prediction calculation | Methods → Analytical methods; Note 1 — p. 6 |
| 13 | Class imbalance | Methods → Class imbalance — p. 7 |
| 14 | Fairness | Methods → Fairness; Table S5 — p. 7 |
| 15 | Model output / thresholds | Methods → Model output and decision thresholds; Table S3 — p. 7 |
| 16 | Training vs evaluation | Methods → Differences between development and evaluation data, including the patient-level separation audit — p. 8 |
| 17 | Ethical approval | Methods → Ethics approval — p. 9 |
| 18a | Funding | Open science (after Conclusion) — p. 23 |
| 18b | Conflicts of interest | Open science (after Conclusion) — p. 23 |
| 18c | Protocol | Open science (after Conclusion) — p. 23 |
| 18d | Registration | Open science (after Conclusion); Abstract → Registration — p. 23 |
| 18e | Data sharing | Open science (after Conclusion) — p. 23 |
| 18f | Code sharing | Open science (after Conclusion) — p. 23 |
| 19 | Patient & public involvement | Methods → Patient and public involvement — p. 23 |
| 20a | Participant flow | Results → Participants and flow — p. 10 |
| 20b | Participant characteristics | Results → Participants and flow; Tables S6, S7 — p. 10 |
| 20c | Development-vs-evaluation distribution | Partially addressed: evaluation partitions are drawn from the same datasets as development (within-distribution), so predictor distributions coincide by construction; no separate comparison was performed. Table S7 characterises the evaluation cohort — p. 8 |
| 21 | n / events per analysis | Results → Model development and specification — p. 10 |
| 22 | Full model specification | Results → Model development and specification (code/weights released) — p. 10 |
| 23a | Performance + subgroups + CIs | Results → Tables 2–5, S5–S10; Figures 2–5, S1–S7. All intervals are patient-clustered BCa bootstrap — pp. 10–18 |
| 23b | Heterogeneity across clusters | Per-cohort performance reported descriptively (Table 2; Table S8); no formal cluster model fitted — pp. 6, 10 |
| 24 | Model updating results | Results → Model updating; Figure S7 — p. 18 |
| 25 | Interpretation | Discussion — pp. 18–20 |
| 26 | Limitations | Discussion → Limitations — pp. 20–21 |
| 27a | Poor/unavailable input handling | Discussion → Usability; Results → Self-reported quality control — pp. 16, 21 |
| 27b | User interaction / expertise | Discussion → Usability in current care and next steps — p. 21 |
| 27c | Future research | Discussion → Usability in current care and next steps — pp. 21–22 |

### Appendix 2. Completed TRIPOD+AI for Abstracts checklist

*Essential items for reporting a prediction-model study in an abstract (Collins GS, et al. BMJ 2024). Reported location within the structured Abstract (manuscript pages 2–3).*

| Item | Topic | Reported in abstract |
| --- | --- | --- |
| 1 | Title | Title (development + evaluation; target population; outcomes) |
| 2 | Background | Abstract → Background |
| 3 | Objectives | Abstract → Objectives |
| 4 | Data sources | Abstract → Methods |
| 5 | Eligibility & setting | Abstract → Methods |
| 6 | Outcome (and time horizon) | Abstract → Methods (concurrent/diagnostic; no time horizon) |
| 7 | Model type / building / internal validation | Abstract → Methods |
| 8 | Performance measures | Abstract → Methods |
| 9 | Number of participants and events | Abstract → Results |
| 10 | Predictors in the model | Abstract → Methods / Results |
| 11 | Performance with confidence intervals | Abstract → Results |
| 12 | Interpretation | Abstract → Conclusions |
| 13 | Registration | Abstract → Registration |
